# From Gut to Brain: Evidence for a Causal Contribution of Gut-Microbiota to Major Depressive Disorder in Humans

**DOI:** 10.1101/2024.12.05.24318549

**Authors:** Leon Fehse, Adèle H. Ribeiro, Nils R. Winter, Sharmili Edwin Thanarajah, Lukas Fisch, Marius Welzel, Corinna Bang, Susanne Meinert, Kira Flinkenflügel, Tiana Borgers, Janik Goltermann, Elisabeth J. Leehr, Mirjam Bloemendaal, Eugenia E. Natasha, Danique Mulder, Alejandro Arias Vasquez, Frederike Stein, Florian Thomas-Odenthal, Paula Usemann, Lea Teutenberg, Igor Nenadic, Benjamin Straube, Nina Alexander, Hamidreza Jamalabadi, Andreas Jansen, Robert Nitsch, Andreas Lügering, Andreas Reif, Sarah Kittel-Schneider, John Cryan, Andre Franke, Benjamin Valderrama, Gerard Clarke, Aonghus Lavelle, Udo Dannlowski, Tilo Kircher, Dominik Heider, Tim Hahn

## Abstract

Major Depressive Disorder (MDD) is a highly prevalent, severe mental health condition that constitutes one of the leading causes of disability worldwide. While recent animal studies suggest a causal role of the gut microbiome in the pathophysiology of MDD models, evidence in humans is still unclear due to small sample sizes, inconsistent clinical assessment of MDD diagnosis, and methodological limitations regarding causal inference in cross-sectional data. Here, we explicitly address these shortcomings to investigate the potential causal link between the gut microbiome and MDD: First, we replicate previous findings using one of the largest multicenter MDD cohorts for which microbiome data and in-depth diagnostic assessment are available (N=1,269 MDD patients and controls). We find a significant difference between healthy controls and MDD patients for the relative abundance of the four taxa *Eggerthella*, *Hungatella*, *Coprobacillus*, and *Lachnospiraceae* FCS020. Second, we employ state-of-the-art, fully data-driven causal inference tools within Judea Pearl’s framework, allowing us to derive model constraints from the data rather than relying on potentially strong, unrealistic assumptions. Using this approach, we found data-driven evidence for *Eggerthella and Hungatella* as causal contributors to MDD. Furthermore, we show that *Eggerthella* and *Hungatella* abundances are associated with MDD beyond the influence of body mass index, identifying two distinct pathways linking MDD to the gut microbiome. Finally, the difference in relative abundance of these taxa between healthy and MDD patients was independent of antidepressant medication. Our study provides the first evidence for a potential causal role of gut-microbiota in the pathophysiology of depression in humans.

## Introduction

Major Depressive Disorder (MDD) is a highly prevalent mental health condition and a leading cause of disability worldwide.^1^ However, the underlying pathophysiology remains poorly understood, and over 30% of patients are unresponsive to current treatments. As neuroimaging research has shown only minimal differences between healthy individuals and MDD patients, a solely neurocentric perspective is challenged.^2–4^ This has shifted attention toward the interplay between the brain and peripheral systems, including the gut microbiome, immune function, endocrine pathways, and metabolism, which may influence neuroinflammation, neurotransmitter synthesis, and stress response.^5^ Understanding these interactions could reveal new pathophysiological insights and therapeutic targets for MDD.

Focusing on the gut microbiome, evidence for a possible role in MDD comes from animal studies employing fecal transplants from depressed patients to microbiota-deficient rats. In this work, rats which had received an MDD fecal transplant consequently showed a depressive phenotype with anhedonia-like (impaired reward response) and anxiety-like behavior.^6^ Rodent studies further indicated that the link between gut-microbiome composition and depression is potentially mediated by modulating neurotransmitter synthesis, peripheral or central inflammation, hypothalamus-pituitary activation and neuronal signaling (see ^7^ for a review). In this context, increased gut permeability with enhanced passage of endotoxins and a dysregulated secretion of pro- and inflammatory metabolites (e.g., short chain fatty acids) by the gut microbiome might play a critical role.

In humans, correlational studies have also linked MDD to the microbiome composition: The systematic review of correlational studies investigating the link between depression and gut-microbiome composition reported reduced abundance of *Faecalibacterium* and increased abundance of *Eggerthella, Atopium* and *Bifidobacterium*. This was corroborated by a recent meta-analysis that found depleted levels of the anti-inflammatory genera *Butyricicoccus, Coprococcus, Faecalibacterium, Fusicatenibacter, Romboutsia* while levels of the pro-inflammatory genera *Eggerthella, Enterococcus, Flavonifractor, Holdemania, Streptococcus* were enriched in MDD patients.^8,9^ While the evidence is encouraging, these investigations solely establish correlations and fail to provide any insight on the causal contribution of microbial taxa to MDD.^10^ Hence, it remains unclear if alterations in the gut-microbiome are the cause or the consequence or an epiphenomen of MDD.

The high prevalence of overweight and obesity, as well as metabolic syndrome in MDD further complicates the relationship, as both conditions are associated with alterations in gut microbiome composition. Such alterations may represent a shared pathway underlying this comorbidity. Notably, fecal transplants from high-fat diet obese mice to lean mice have induced depressive-like behaviors, indicating that gut microbiome changes contribute to the depression-obesity link.^11^ Previous research either ignored the impact of body mass index (BMI) or included it as a covariate in the study without further examining its differential impact on MDD and gut microbiota composition.

From a methodological point of view, causally contributing microbial genera as therapeutic targets for MDD has faced three main challenges: (1) most human studies have small samples (n<70; see ^8^ for a systematic review), limiting statistical power, increasing false positives, and reducing generalizability, (2) inconsistent clinical assessments of MDD, often based on self-reported questionnaires rather than clinician-led interviews, result in unreliable diagnoses and varied patient samples^12^, (3) most studies are only cross-sectional but not longitudinal, hampering the establishment of causal relationships (4) causality inferred from genetic instrumental variable techniques, such as Mendelian randomization^13,14^ and certain machine learning-based causal inference methods^15^ may rely on unrealistic assumptions concerning the roles of hidden confounders (see, e.g.,^16,17^).

To overcome these limitations, we replicated previous findings using one of the largest multicenter MDD cohorts for which microbiome data and in-depth diagnostic clinical assessment is available (N=1,269 MDD patients and controls). Methodological heterogeneity was minimized in this well-curated, harmonized sample^18^. We employed state-of-the-art computational approaches for causal inference within Judea Pearl’s structural framework of causation^19,20^. This framework represents a seminal advancement in data analysis, enabling us to move beyond detecting mere statistical associations to identifying causal relationships directly from observational data. Although based on data-driven statistical associations and independencies, the notion of causality inferred by this framework aligns with the interpretation derived from randomized controlled experiments: a variable X is considered a cause of another variable Y if an intervention on X leads to an expected change in Y. While randomized experiments rely on perfect randomization of treatment assignment to minimize confounding effects, this framework infers causality by systematically identifying and adjusting for the potential influence of confounders, including those that may be unobserved. Note that, as with randomized controlled experiments, determining that X causes Y does not imply that X is the sole cause of Y or that the effect is not mediated by other unobserved causal factors.

In our causal data analysis, we specifically employ causal discovery algorithms to effectively learn a class of graphical causal models that fit the observed data while accounting for potential unobserved confounders^20^. We then apply effect identification and estimation tools to infer unbiased causal effects, free from the influence of (observed and unobserved) confounders, based on the learned causal structure, in a fully data-driven manner^21,22^. Notably, our approach offers significant advantages over causal inference methods that rely on strong, often unrealistic assumptions, such as unconfoundedness (i.e., the set of observed covariates suffices for confounding adjustment) in most causal machine learning-based approaches^15^ or valid instruments in Mendelian Randomization. Due to numerous issues such as unobserved confounders, bias-amplifying variables, M-bias^23,24^, and horizontal pleiotropy^16,17^, these assumptions are very likely to be violated in practice, making reliable causal inferences challenging without rigorous methods.

## Methods

### Study Design and Participants

Study participants are part of the Marburg-Münster Affective Disorders Cohort Study (MACS).^13^ Data were collected at two sites (Marburg and Münster, Germany) using identical study protocols. N=2,035 healthy participants and patients with MDD were recruited as part of the MACS cohort from September 11, 2014, to September 26, 2018. MDD diagnosis was assessed using the Structured Clinical Interview for DSM-IV, axis 1 disorders (SCID-I)^25^. Patients either fulfilled the DSM-IV criteria for an acute major depressive episode or had a lifetime history of a major depressive episode. Individuals with any history of neurological or medical conditions were excluded, resulting in a sample of N=1,802. Fecal samples were available for N=1,313 individuals. From these, 44 samples were excluded because of missing values for BMI, resulting in a sample size of N=1,269 samples (644 HC, 625 MDD). For further details on the exclusion criteria see supplementary materials.

The FOR2107 cohort project was approved by the Ethics Committees of the Medical Faculties, University of Marburg (AZ: 07/14) and University of Münster (AZ: 2014-422-b-S). Participants received financial compensation and gave written and informed consent.

### Assessment of Symptom Severity

#### Hamilton Depression Score (HAM-D)

A widely employed clinician-administered tool to assess depression severity. The scale consists of 21 items, each rated on a scale from 0 to 4 or 0 to 2, depending on the item. Higher scores indicate a greater severity of depressive symptoms.

#### Beck Depression Inventory (BDI-I)

This is a questionnaire to assess self-reported depressive symptoms. The 21-item questionnaire is designed to capture the intensity of depressive symptoms based on the respondent’s experiences over the past week. Each item is rated on a scale from 0 to 3, with higher scores reflecting greater severity of depressive symptoms. The total score is calculated by summing the responses, with higher scores indicating a higher level of depressive symptomatology.

### Fecal Sample Collection

The dataset consists of 1,269 standardized human stool samples of which 16S rRNA has been sequenced. For homogeneity, all samples were collected following a standardized protocol for sampling and analysis.

### DNA Extraction, PCR, and Sequencing

The procedure for DNA extraction and sequencing has been described previously.^26,27^ DNA was extracted from fecal samples using the QIAamp DNA fast stool mini kit automated on the QIAcube (Qiagen, Hilden, Germany). Therefore, material was transferred to 0.70 mm Garnet Bead tubes (Dianova, Hamburg, Germany) filled with 1. 1  InhibitEx lysis buffer. Bead beating was performed using a SpeedMill PLUS (Analytik Jena, Jena, Germany) for 45 s at 50 Hz. Samples were then heated to 95°C for 5 min with subsequent continuation of the manufacturer’s protocol. Extracted DNA was stored at −20°C prior to PCR amplification. Blank extraction controls were included during extraction of samples.

For sequencing, variable regions V1 and V2 of the 16S rRNA gene within the DNA samples were amplified using the primer pair 27F-338R in a dual-barcoding approach according to Caporaso et al. (PMID: 22402401).^28^ 3 µ stool DNA were used for amplification. PCR-products were verified using the electrophoresis in agarose gel. PCR products were normalized using the SequalPrep Normalization Plate Kit (Thermo Fisher Scientific, Waltham, MA, USA), pooled equimolarity and sequenced on the Illumina MiSeq v3 2×300bp (Illumina Inc., San Diego, CA, USA). Demultiplexing after sequencing was based on 0 mismatches in the barcode sequences.

### Preprocessing of Sequencing Data

To process raw sequencing data, identify operational taxonomic units (OTU’s) in the 16S rRNA data and find the abundances of corresponding taxonomies, we used the amplicon read processing pipeline Natrix.^29^ Natrix combines quality control, read assembly, primer removal, dereplication, chimera detection, abundance filtering, OTU generation, and assignment of taxonomic information into a single workflow using the rule-based Snakemake workflow management engine.^30^ Low-quality reads were filtered using PRINSEQ^31^ (v0.20.4) at a PHRED score threshold of 30. The primer removal and assembly of forward- and reverse-reads were carried out using the PANDAseq paired-end assembler.^32^ CD-HIT-EST^33^ was used for the dereplication, by clustering sequences with 100% identity. Chimera detection and removal was carried out via the uchime3_*denovo* algorithm of the VSEARCH^34^ framework. The SWARM clustering algorithm^35^ was used to identify OTUs, and the assignment of taxonomic information to the OTUs was carried out using BLAST^36^ with the curated SILVA rRNA database.^37^

### Microbiome association analysis

We used Natrix to generate the taxonomic profiles as described above. OTUs for which Natrix could not assign taxonomy on genus level as well as samples lacking relevant metadata were filtered out. Furthermore, the data was updated to use current scientific naming used by NCBI and formatted to fit the broad applicability in subsequent general statistical analysis and differential abundance analysis (DAA). These statistical evaluations were conducted utilizing the R programming language, specifically employing the microbiome analysis package *mia* (v. 1.10.0) for data loading. Prior to the DAA, genera with a prevalence of less than 10% (i.e., with non-zero values in less than 10% of the samples) were removed from the data.

### Alpha and Beta Diversity

Alpha and beta diversity are metrics describing the difference of diversity within samples (alpha) and between samples (beta) and are typically reported in microbiome related studies. Logistic regression analysis was carried out to examine the association between MDD and various descriptive variables. In this analysis, MDD served as the target variable, while sex, age, body mass index (BMI), library size (the sum of all read counts in a sample), site (which declares whether the data was collected in Marburg or Muenster) and alpha diversity, represented by the Shannon index, were used as predictors.

To cover beta diversity a Permutational Multivariate Analysis of Variance (PERMANOVA) was conducted on Bray-Curtis dissimilarity using the dissimilarity as the target and MDD, along with sex, age, BMI, site and library size as predictors, to assess the connection between beta diversity and MDD. This was done using the *adonis3* function from the *GUniFrac* package (v. 1.8).

### Differential Abundance Analysis

We carried out a DAA to gain insights into the roles of various taxa within the human gut microbiome in relation to MDD. To this end, OTUs were grouped at the genus level and counted. The analyses were carried out on genus level due to the known limitations of amplicon sequencing methods, particularly the 16S gene sequencing used in this study, which does not provide species-level resolution.^38^ The top 3 percentiles of each genus’ values were identified as outliers.

Since microbiome data typically exhibits high variance in read counts, compositional effects, and zero-inflation, specialized models are required to analyze taxa. Therefore, we employed two different models, namely ZicoSeq^39^ and LinDA^40^, implemented in the GUniFrac (v. 1.8) and MicrobiomeStat (v. 1.1) packages, respectively. ZicoSeq utilizes a linear regression approach on log-transformed taxa abundance values against specific covariates, correcting for compositional effects using a reference-based strategy and an empirical Bayes method to handle zero-inflation. LinDA similarly fits a linear regression model on log2-ratio-transformed taxonomic abundance data, also accounting for covariates, but addresses compositional effects and zero-inflation through sample library size adjustments. Both models apply winsorization to manage outliers in taxonomic abundance data.

These analyses produced p-values indicating the significance of the relationship between each genus and MDD. Within the statistical models, we controlled for age, sex, BMI, site and library size as covariates. To adjust for multiple testing, we used false discovery rate (FDR) correction. Specifically, ZicoSeq employed a permutation-based FDR control, whereas LinDA utilized the Benjamini-Hochberg (BH) procedure. Genera that remained significant at a significance level of 0.05 were then subjected to further downstream analyses investigating the causal relationship between genera and MDD.

### Causal Analyses

#### Identifying Potential Causal Relationships between Microbial Taxa and MDD

To uncover causal relationships between MDD and microbial taxa significantly associated with MDD, we run a causal discovery analysis using Fast Causal Inference (FCI)^41^. FCI is distinguished by its rigorous foundational principles and minimal reliance on assumptions compared to other causal discovery methods. When integrated with Zhang’s^20^ orientation rules, FCI can provide reliable and complete results, even in the presence of unobserved confounding and selection bias. Additionally, the FCI is sound and complete even when feedback loops are present (i.e., the underlying model is a dynamical system in equilibrium)^42^. This ensures that, whenever statistical associations are accurately inferred, the FCI output reliably estimates both the presence and absence of ancestral (causal) relationships, as well as the absence of confounders and cycles. The FCI algorithm is available in the *pcalg* R package.^43^ The covariates sex, age, library size, BMI, and site are integrated into the analysis.

For detailed information on the FCI algorithm, see the supplementary materials. In short, the FCI algorithm starts with an adjacency phase, where multiple conditional independence tests are performed to learn the *skeleton* of the underlying causal model – an undirected graph representing potentially direct associations among the observed variables. Then, it proceeds to the orientation phase, where a set of rules are applied to determine the directionality of edges. These rules identify all causal relationships (arrow tails) and non-causal associations (arrow heads) that can be inferred from observational data. This results in a Partial Ancestral Graph (PAG) representing the class of statistically equivalent models known as Markov Equivalence Class (MEC). In a PAG, relationships that are not uniquely determined due to equivalent structures are represented by circle edge marks.

As our dataset includes both binary and continuous variables, we use the symmetric conditional independence test for mixed data proposed by Tsagris et al.^44^, available in the MXM R package^45^. For testing conditional independencies involving microbial taxa, we extended Tsagris et al.’s^44^ approach to use LinDA and ZicoSeq.

To ensure reliability, we implement several robustness assessments. First, we evaluate marginal causal consistency following the methodology outlined by Roumpelaki et al.^46^. This involves applying the FCI algorithm to every possible subset of our selected variables, aiming to evaluate the consistency of inferred causal relationships across all marginal analyses. Additionally, we validate each inferred PAG to ensure it conforms to the expected characterization and implies only conditional dependencies and independencies consistent with our observed data. Instances revealing violations or inconsistencies are systematically excluded from our analysis. Finally, we compare all PAGs that passed our validity and robustness tests with those obtained using the conservative variant of the FCI algorithm^47^. This variant employs additional tests to identify ambiguous triplets and adopts a conservative approach to edge orientations, relying exclusively on triplets previously identified as unambiguous.

### Causal Effect Estimation

A PAG offers a qualitative representation of the class of all causal models compatible with the observed data, with directed edges denoting causal relationships. To quantitatively assess the causal effect that one variable has on another, it is essential to conduct an effect identification analysis. The identification of a causal effect is contingent upon its uniqueness -- it is identifiable if and only if it is uniquely computable among all models within the equivalence class represented by the PAG, and utilizing the same expression solely based on observational (conditional) probabilities.

Several methods and algorithms exist for effect identification in PAGs. These include the generalized backdoor criterion^48^, the generalized adjustment criterion^21^, and the IDP and CIDP algorithms for identifying (conditional) causal effects^22^. Notably, IDP and CIDP are the only complete effect identification algorithms for PAGs. This means they can identify every (conditional) causal effect that is possible to be identifiable, possibly through methods distinct from backdoor or adjustment criteria. This makes them particularly powerful tools in causal analysis. Additional details on the identification and estimation of the causal effects can be found in the supplementary materials.

### Medication Effects

For the taxa identified as causal contributors to MDD, we further tested whether the use of psychopharmacological medication could account for differences in their relative abundance between MDD patients and healthy controls. For this purpose, we stratified the patient group by medication status. Patients taking at least one psychopharmacological medication (e.g. antidepressants, antipsychotics, anti-epileptics, see Supplementary Methods 4 for detailed information) were categorized as medicated, while patients without any psychopharmacological medication were classified as unmedicated. We performed logistic regression to test for differences in CLR-transformed relative abundance between the medication groups, as well as between each of the stratified patients groups and the unaffected controls. All models were adjusted for age, sex, BMI, collection site, and library size. We performed Benjamini-Hochberg (BH) procedure on the p-values to correct for multiple comparisons.

## Results

We present a comprehensive analysis investigating the role of the gut microbiome in MDD. First, aiming to replicate findings reported in the literature, we explore the associations of alpha and beta diversity with MDD, followed by a differential abundance analysis to identify specific microbial taxa linked to the disorder. Based on the taxa significantly associated with MDD, we subsequently utilize causal inference methods to identify and quantify the causal relationships between microbial taxa and MDD, further assessing the causal effect of microbial taxa on MDD across BMI categories.

### Sample Characteristics

Table 1 provides key descriptive statistics for the MACS cohort, encompassing a total of 1,269 individuals. Of these, 644 are healthy controls (HC) (213 male; 412 female), and 625 are patients with MDD (217 male patients; 427 female patients). Our logistic regression analyses revealed no significant differences between the two groups in terms of male/female ratio (p=0.7394), age (p=0.0728), library size (p=0.5901), and alpha diversity (Shannon Index) (p=0.3474). However, significant differences were observed between the two groups for BMI and site (p<0.001). For the full results of this analysis see *Supplementary Table 1*.

**Table 1:** Descriptive statistics for all samples used in the analysis divided into the groups MDD and HC.

|  | Healthy controls | MDD patients | P-values |
| --- | --- | --- | --- |
| Total Number of Samples | 644 | 625 |  |
| Sex (Number of Samples) | Male: 217; Female: 427 | Male: 213; Female: 412 | 0.739 |
| Site (Number of Samples) | Münster: 366; Marburg: 278 | Münster: 427; Marburg: 198 | <0.001 |
| Age (Mean ± SD) | 35.73 ± 13.32 | 36.98 ± 13.24 | 0.073 |
| BMI (Mean ± SD) | 24.36 ± 4.65 | 26.13 ± 5.86 | <0.001 |
| Library Size (Mean ± SD) | 19,968.79 ± 7,538.20 | 20,958.16 ± 9,184.63 | 0.590 |
| Shannon Index (Mean ± SD) | 2.77 ± 0.329 | 2.745 ± 0.336 | 0.347 |
| BDI | 4.10 ± 4.30 | 16.89 ± 11.04 | <0.001 |
| HAMD | 1.60 ± 2.33 | 9.28 ± 7.27 | <0.001 |
| Remission status |  | Acute: 417; Remitted: 208 |  |
| Psychopharmacological Medication |  | Unmedicated: 246; Medicated: 379 |  |

## Main Results

### No difference in alpha and beta-diversity between MDD patients and healthy controls

To understand the overall composition and diversity of the gut microbiome in relation to MDD, we conducted analyses of both alpha and beta diversity. Testing significance in alpha diversity with a logistic regression based on individual Shannon index values did not indicate a significant difference between MDD and HC samples (p=0.3570). Similarly, beta diversity assessed via PERMANOVA on individual Bray-Curtis dissimilarity values did not reveal significant differences between the two groups (R²<0.001, p=0.122).

### Differential Abundance Analysis revealed four taxa that differed between MDD patients and healthy controls

To identify specific microbial genera associated with MDD, we conducted a Differential Abundance Analysis (DAA). Following data preprocessing, our dataset included 28,461 Operational Taxonomic Units (OTUs) across all samples. For the DAA, we filtered these OTUs based on their prevalence at the genus level, resulting in 172 distinct genera.

For the genus-level DAA, we used two models: ZicoSeq and LinDA. The ZicoSeq model identified four genera that significantly differed between MDD and HC individuals: *Eggerthella*, *Hungatella, Lachnospiraceae FCS020 group*, and *Coprobacillus* (see *Supplementary Figure 1*). The LinDA model identified seven significantly different genera: *Eggerthella*, *Hungatella*, *Clostridium sensu stricto 1*, *Faecalibacterium*, *Coprobacillus*, *Lachnospiraceae FCS020 group*, and *Subdoligranulum* (see *Supplementary Figure 2*).

We focused further causal analysis on the set of seven genera identified by LinDA, with special emphasis on *Eggerthella*, *Hungatella*, *Coprobacillus*, and *Lachnospiraceae FCS020 group*, as these were consistently identified by both the ZicoSeq and LinDA models. Adjusted *p*-values for these four genera in both models are provided in the *Supplementary Table 3*.

### Causal Analyses identified *Eggerthella* and *Hungatella* as causal contributors to MDD

The set of genera found significantly associated with MDD by both ZicoSeq and LinDA, namely *Eggerthella*, *Hungatella, Coprobacillus,* and *Lachnospiraceae FCS020 group*, underwent further causal analysis. Our goal was to uncover causal relationships among MDD and these four genera, while also considering the influence of the observed potential confounders. Our strategy involved employing the FCI algorithm to recover the graphical model over these 5 variables, alongside BMI and site. Furthermore, to ensure that all relationships are controlled for age, sex, and library size, these crucial known confounders were incorporated as a fixed component of the conditioning set for all conducted conditional independence tests.

As detailed in the methods section, to ensure reliability of our findings, we systematically ran the FCI across all feasible subsets of the selected variables, each consisting of a minimum of 3 variables, resulting in a total of 31 PAGs. We exclusively focused on the resulting PAGs that passed our validity and robustness tests, i.e., those with no conflicting orientations across marginals, no inconsistencies in the implied conditional independencies, and no violations of the inherent PAG properties. Then, we assessed the orientations within those PAGs that were inferred identically by both the FCI and its conservative variant. We executed this approach twice, once using ZicoSeq and again using LinDA to test for conditional independencies involving microbial taxa variables. *Supplementary Table 4* summarizes the orientations consistently inferred using both the FCI and conservative FCI algorithms while using LinDA, and *Supplementary Table 5* summarizes those obtained using ZicoSeq.

While using conditional independence tests based on LinDA, the following definite relationships were robustly identified, with no ambiguity or conflicting orientations: *Eggerthella* and *Hungatella* both cause MDD (2 PAGs), *Coprobacillus* causes *Eggerthella* (8 PAGs), the *Lachnospiraceae FCS020 group* is only spuriously associated with both *Coprobacillus* and *Eggerthella* (2 PAGs), and *Hungatella* is also only spuriously associated with both *Coprobacillus* and *Eggerthella* (6 PAGs). Moreover, the following definite non-ancestral relationships were obtained: MDD is not an ancestor (cause) of BMI (20 PAGs), Site (18 PAGs), *Lachnospiraceae FCS020* group (12 PAGs). Note that a definite non-ancestral relationship leaves open the possibility of causality in the opposite direction or purely spurious associations, as they could not be conclusively confirmed with the current data. The remaining valid PAGs identified consistent, although less informative orientations. While using ZicoSeq, only definite non-ancestral relationships were obtained robustly, including that MDD is not an ancestor (cause) of BMI (11 PAGs), Site (10 PAGs), *Coprobacillus* (4 PAGs), and *Eggerthella* (4 PAGs). Significantly, while less informative, those and all other PAGs inferred using conditional independence tests based on ZicoSeq are consistent with those obtained using those based on LinDA.

The PAG in Figure 1 shows all robustly identified causal relationships between taxa variables and MDD. It indicates that *Hungatella* and *Eggerthella* are causes of MDD, and that *Coprobacillus* causes MDD through *Eggerthella*. Importantly, these causal relationships are identified as unconfounded by latent variables and have no cycles. BMI is identified as a confounder of the relationships among the microbial taxa variables and MDD. The PAG obtained by the conservative FCI, shown in the Supplementary Material, supports these causal and confounded relationships but shows uncertainty in the causal link between BMI and MDD due to ambiguity in the *Coprobacillus*-BMI-MDD triplet. None of the valid PAGs identified a definite relationship between the *Lachnospiraceae* FCS020 group and MDD.

**Figure 1:**
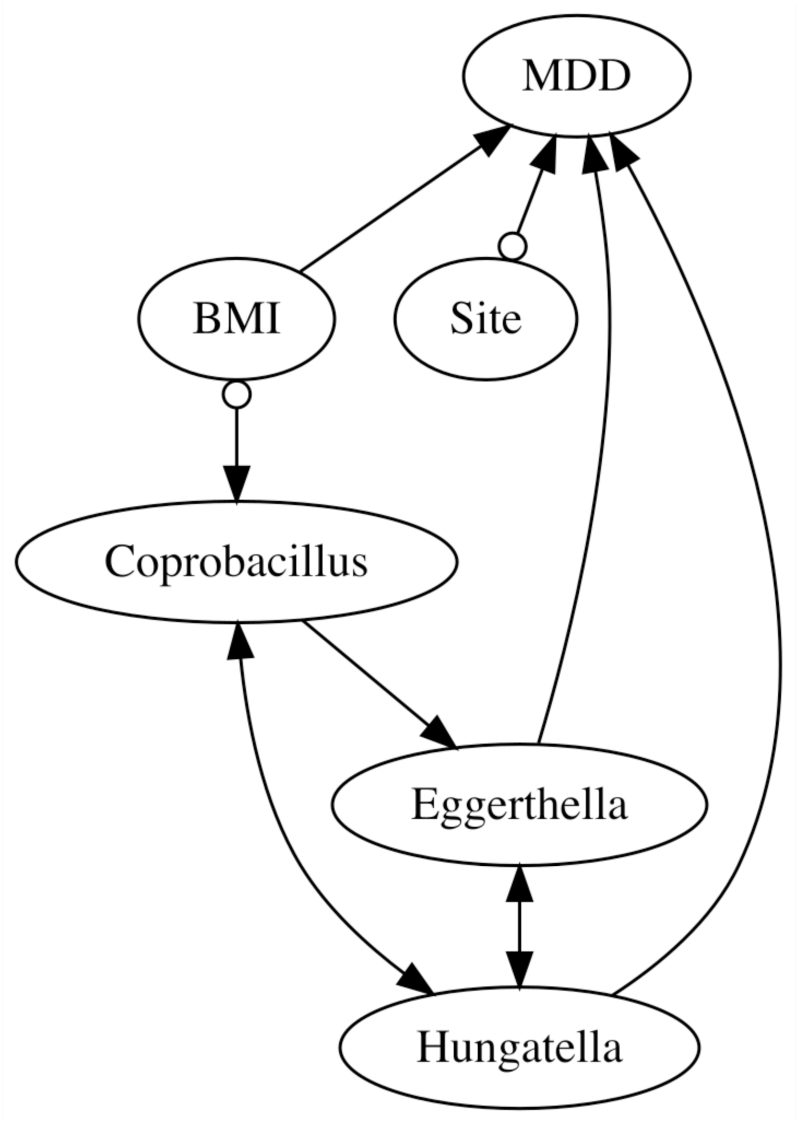
FCI’s resulting PAG over MDD, BMI, Site and all microbial taxa robustly identified as causes of MDD, namely *Eggertella*, *Hungatella*, and *Copobracillus*. Conditional independence tests for taxa variables rely on LinDA and consistently include age, sex, and library size as part of the conditioning set. A directed edge indicates an ancestral (causal) relationship. A bidirected edge indicates a non-causal relationship, due to only latent confounders. Circles indicate non-definite (uncertain) relationships.

### The causal effect of *Eggathella* and *Hungatella* on MDD was strong across all BMI groups

Figure 2 illustrates the causal effect of *Eggerthella* and *Hungatella* on MDD, expressed as the probability of experiencing MDD given a specific abundance level "e" of Eggerthella set by intervention (P(MDD=1|do(*Eggerthella*=e))), and abundance level "h" of *Hungatella* set by intervention (P(MDD=1|do(*Hungatella*=h))), respectively. The probability of MDD significantly increases with higher levels of *Eggerthella* or *Hungatella*. For example, when the abundance of *Eggerthella* is set to 0, the probability of MDD is 47.2% (95% CI: 42.6% to 52.0%). This probability rises to 60.8% (95% CI: 52.2% to 68.8%) when the abundance is set to 70. When *Hungatella* abundance is 0, the probability of MDD is 47.4% (95% CI: 42.8% to 52.1%), increasing to 53.9% (95% CI: 53.9% to 70.2%) when the abundance is set to 11. These effects were identified also by controlling for both BMI and *Hungatella* and BMI and *Eggerthella*, respectively. Further details on the identification and estimation procedures are provided in the Supplementary Material.

**Figure 2:**
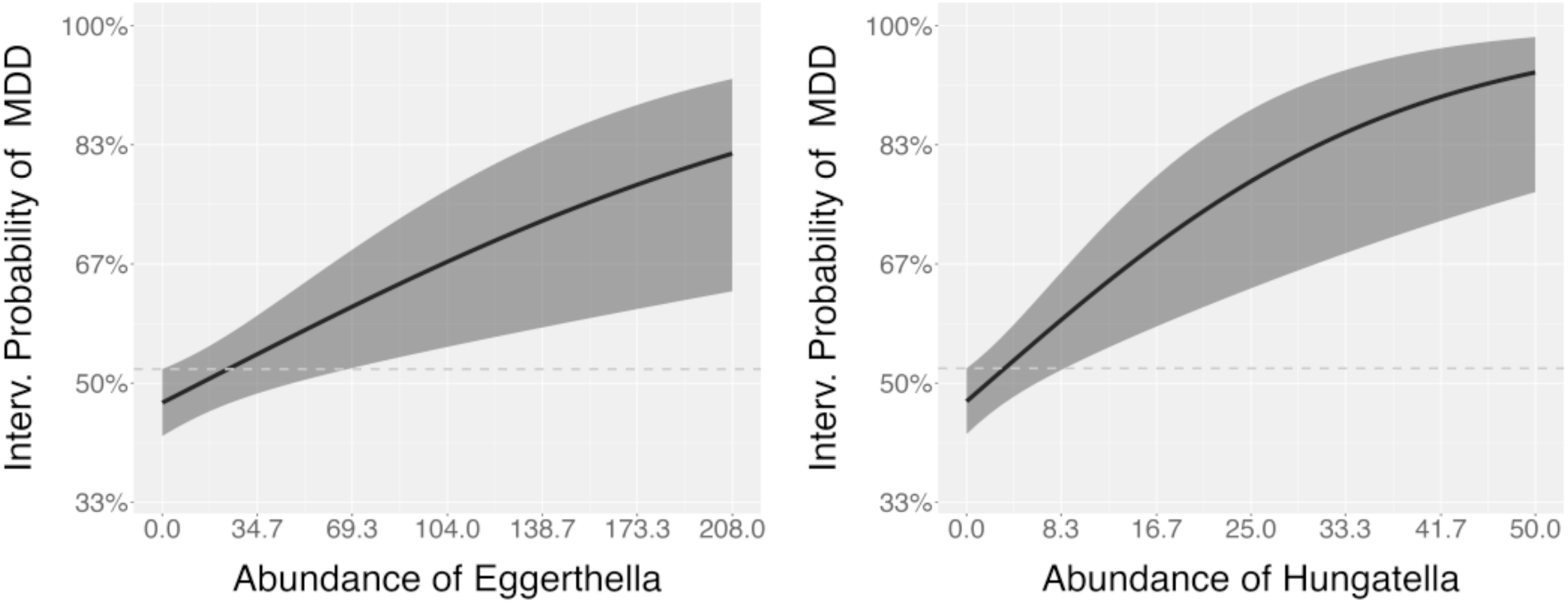
Estimated post-interventional probabilities of MDD and respective 95% confidence intervals given (left) *Eggerthella* abundance levels (counts) "e", P(MDD=1∣do(Eggerthella=e)), and (right) *Hungatella* abundance levels (counts) "h", P(MDD=1∣do(Hungatella=h)).

We further analyzed the causal effect of *Eggerthella* and *Hungatella* on MDD across BMI groups. BMI values were categorized into three groups: "Non-Obese" (BMI 14-25), "Overweight" (BMI 25-30), and "Obese" (BMI >30). The dataset includes 746 non-obese individuals, 334 in overweight, and 189 obese. Subsequently, we assessed the causal effects of *Eggerthella* and *Hungatella* on MDD within each of these BMI categories.

Figure 3 shows the causal effect of *Eggerthella* on MDD in each category of BMI "c", expressed as the post-interventional probability distribution P(MDD=1|do(*Eggerthella*=e), catBMI=c), alongside the corresponding 95% confidence region. The analysis reveals a significant increase in the probability of MDD with increased *Eggerthella* abundance levels, regardless of the individual’s BMI category. Similarly, Figure 4 illustrates the causal effect of *Hungatella* on MDD, conditional on BMI category "c", expressed as P(MDD=1|do(*Hungatella*=h), catBMI=c), with the corresponding 95% confidence region. Again, there is a significant increase in the probability of MDD with higher *Hungatella* abundance levels, regardless of the individual’s BMI category.

**Figure 3:**
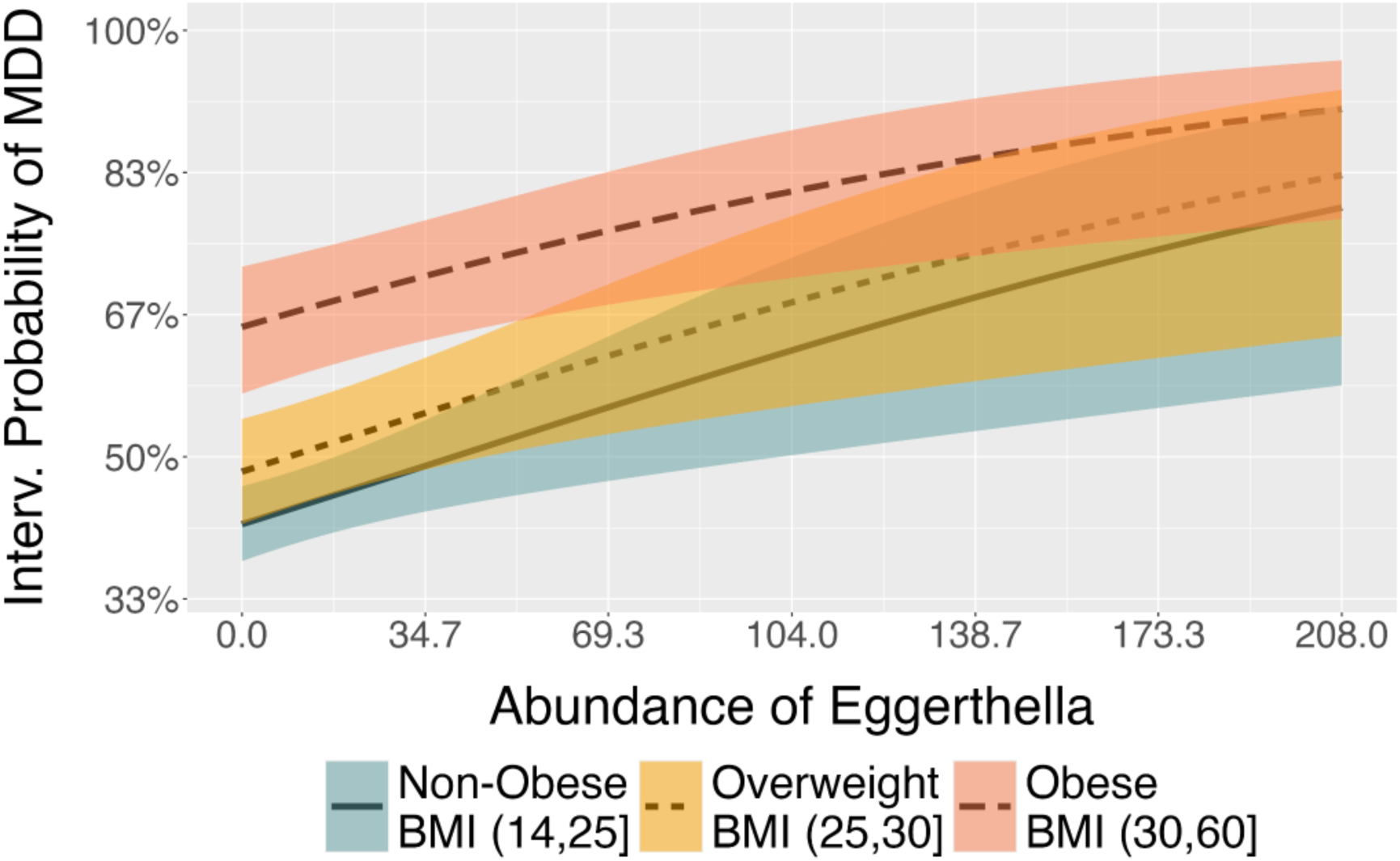
Estimated values of P(MDD=1|do(*Eggerthella*=e), catBMI=c), i.e., the post-interventional probabilities of MDD given that *Eggerthella* abundance values "e" are set by intervention, for each category of BMI "c", along with the respective 95% confidence regions.

**Figure 4:**
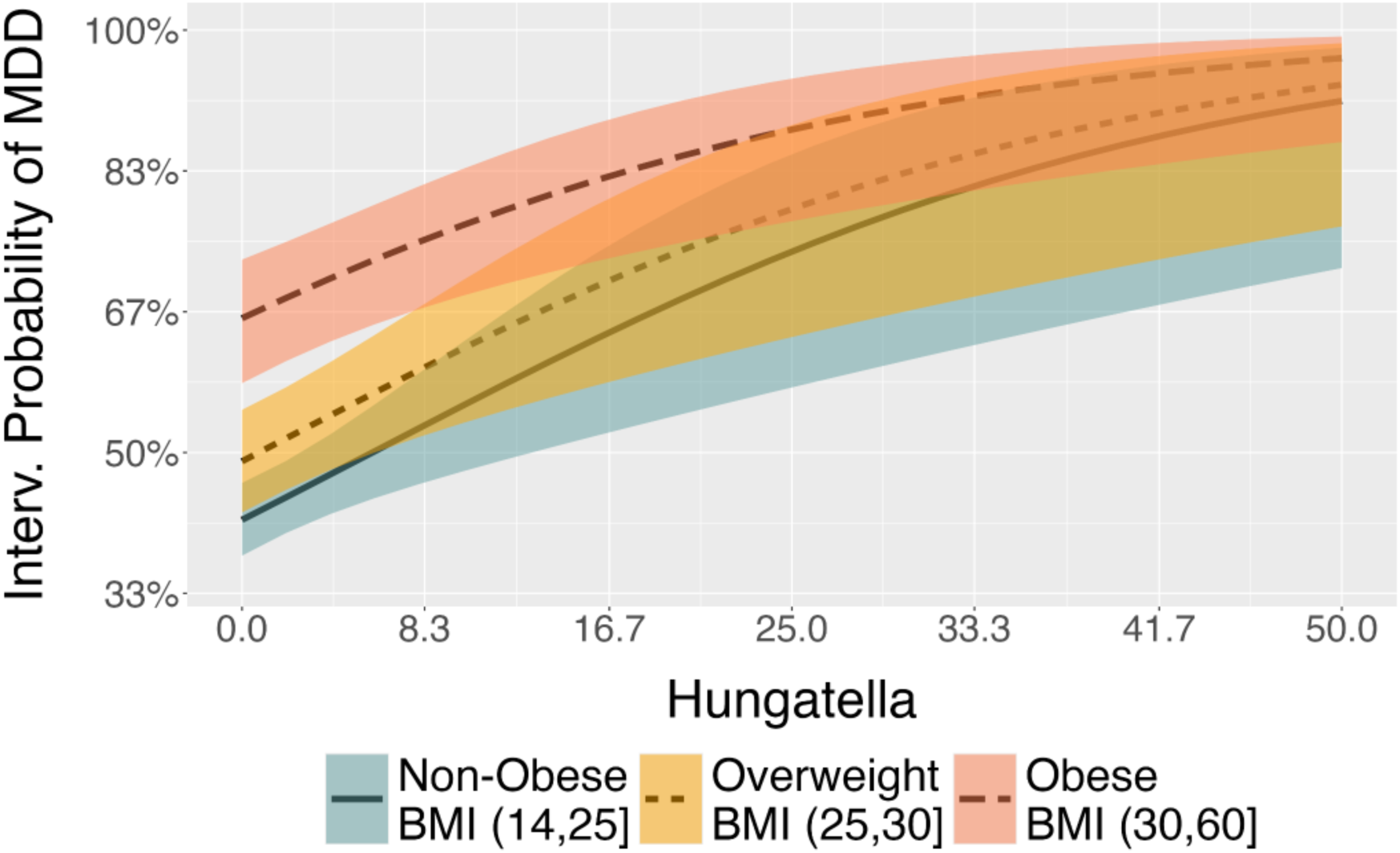
Estimated values of P(MDD=1|do(*Hungatella*=h), catBMI=c)), i.e. the post-interventional probabilities of MDD given that *Hungatella* abundance values "h" are set by intervention, for each category of BMI "c", along with the respective 95% confidence regions.

### The contribution of *Eggerthella* and *Hungatella* to MDD is independent of psychopharmacological medication

Finally, we tested whether the relative abundance of *Eggerthella* and *Hungatella* depended on medication. We distinguished participants that were completely unmedicated (N=244) and participants that were on any kind of psychopharmacological medication (N=360, please find detailed information in Supplementary Method 4). Compared to the healthy controls (N=626), both unmedicated and medicated MDD patients had a higher relative abundance of *Eggerthella* (q=0.04 and q=0.001) and *Hungatella* (q=0.002 and q<0.001), while there was no significant difference between unmedicated and medicated MDD patients (q=0.248 and q=0.282, respectively), indicating that the association between MDD diagnosis and relative abundances of *Eggerthella* and *Hungatella* was independent of medication use (see also *Supplementary Table 2*).

## Discussion

Our study provides evidence for a potential causal role of the gut microbiome in the pathophysiology of major depressive disorder (MDD) using one of the largest multicenter cohorts of clinically well-characterized MDD patients and healthy controls to date. We identified significant differences in the relative abundance of four bacterial taxa—*Eggerthella, Hungatella, Coprobacillus, and Lachnospiraceae FCS020*—between MDD patients and healthy controls. Notably, *Eggerthella* and *Hungatella* were identified as causal contributors to MDD using state-of-the-art, fully data-driven causal inference tools within Judea Pearl’s Structural Causal Model framework. These findings were consistent when accounting for the influence of body mass index (BMI), medication indicating that the relationship between these microbial taxa and MDD go beyond obesity-related factors.

### Gut microbiota causally contribute to depression

Our findings extend previous association studies reporting a clear link between depression and gut-microbiome composition. While in line with our findings most of the studies could not find differences on indices of alpha- or beta-diversity between MDD-patients and healthy controls^8^, differences were observed on the levels of bacterial genera. Particularly depleted levels of the anti-inflammatory genera *Butyricicoccus, Coprococcus, Faecalibacterium, Fusicatenibacter, Romboutsia* and increased levels of the pro-inflammatory genera *Eggerthella, Enterococcus, Flavonifractor, Holdemania, Streptococcus* were found in MDD patients in recent meta-analyses.^8,9^ While the evidence is encouraging, these studies were largely based on small sample sizes (according to ^8^, the mean number of included MDD patients was 48) or quantified depression by self report questionnaires in population based cohorts not requiring a clinical diagnosis.^10^ Hence, the contribution of gut microbiota alterations to clinically diagnosed MDD remained unclear and it is largely unexplored whether alterations in the gut-microbiota are the cause of MDD or merely a consequence, potentially due the secondary effects of antidepressants or comorbid obesity.

Using state-of-the art inference tools, we addressed this limitation, and demonstrate that among the four bacterial genera - *Eggerthella, Hungatella, Coprobacillus, and Lachnospiraceae FCS020* - differing in their relative abundance between MDD patients and healthy controls in our sample, *Eggerthela* and *Hungatella* were causal contributors to MDD. These findings extends previous fecal transplant studies, where rats receiving fecal transplants from MDD patients developed a depressive phenotype^6^, while the symptoms were alleviated after fecal transplants from healthy donors indicating a causal role of the microbiome, while the contribution of single taxa remained unclear.^49^ Notably, a recent Mendelian randomization analysis in the population based Rotterdam cohort, identified a causal contribution of *Eggerthella* to depressive symptoms in humans. It is important to acknowledge, however, that depressive symptoms in this cohort were identified by a self-report questionnaire and did not require a clinical diagnosis.^12^

The use of modern, data-driven causal inference tools within Judea Pearl’s structural framework represents a significant methodological advancement in this study. Unlike other methods that rely on predefined causal models with potentially restrictive assumptions, the methods employed here derive all model constraints directly from data using causal discovery techniques. This shift towards data-driven causal inference not only enhances transparency of the underlying causal structure but also ensures the robustness and interpretability needed to identify potential therapeutic targets.

### The relevance of *Eggerthella* and *Hungatella* in the pathophysiology of depression

Both *Eggerthella* and *Hungatella* are pro-inflammatory taxa that are only present in low levels in the gut-microbiota of healthy adults and adolescents (see *Supplementary Figure 1*). *Eggerthella lenta*, a gram-positive, anaerobic species, has been repeatedly associated with MDD, as well as other chronic (auto-immune) conditions such as multiple sclerosis^50^, rheumatoid arthritis, and asthma bronchiale^51^ highlighting its potential contribution to systemic inflammations. Interestingly, a recent study using a humanized rodent model of rheumatoid arthritis demonstrated that colonization with *Eggerthella* induced a change in the gut-microbiota composition with a pro-inflammatory metabolic-shift with reduced fecal levels of the short-chain fatty acid butyrate, increased levels of acetate and elevated serum cytokine levels.^52^ This finding is highly relevant given increasing evidence, that butyrate is reduced in MDD^53,54^ and chronic low-grade inflammation plays a relevant role in the pathophysiology of depression at least in a subgroup of patients termed “immuno-metabolic depression”.^55^ Moreover, *Eggerthella* gavage in rodents reduced the serum levels of tryptophan^52^, that is the precursor of serotonin, the key neurotransmitter targeted by antidepressant medication Notably, a recent transdiagnostic meta-analysis across multiple psychiatric conditions, including anxiety disorder, major depressive disorder (MDD), bipolar disorder, psychosis, and schizophrenia provided most consistent evidence for the increased relative abundance of *Eggerthella* (reported in 10 out of 11 studies) suggesting that elevated *Eggerthella* levels may be part of a shared pathophysiological pathway that increases vulnerability to mental health impairments.^9^ *Hungatella* has also been linked to MDD in previous studies, though the evidence is less robust compared to the association with *Eggerthella.*^12^ Interestingly, in our study the effect of *Hungatella* on MDD was higher than for Eggerthella (19.84% vs 16.83%). *Hungatella hathewayi* are gram-positive, anaerobic bacteria.^56^ Increased relative abundance of *Hungatella* has been associated with different disorders including eczema in children^57^ and different cancer entities.^58^ *Hungatella* was also associated with impaired cognitive performance and increased gut-permeability quantified by zonulin levels in a sample of Schizophrenia patients.^59^ These findings clearly underscore the relevance of *Hungatella* and *Eggerthella* in the pathophysiology of MDD, while future studies need to elucidate the exact mechanisms.

Exploring the association of changes in gut-microbiota and depression it is important to account for the impact of BMI. Obesity is one of the most common somatic comorbidities^60^ and is linked to changes in the the gut-microbiota.^61^ Considering BMI and overweight/obesity categories, we could demonstrate that the causal contribution of relative *Eggerthella* and *Hungatella* abundance to MDD extends beyond their impact on BMI.

### Increased abundance of *Hungatella* and *Eggerthela* in MDD is independent of psychiatric medication use

Another key factor influencing the gut microbiota is medication, with antidepressants in particular shown to affect its balance and diversity.^62,63^ Interestingly, previous meta-analyses comparing medicated and unmedicated patient groups did not find a significant impact of antidepressants on *Eggerthella* or *Hungathell*a. This aligns with our findings, which show no differences in the relative abundance of these taxa between medicated and unmedicated patients, while both MDD groups significantly differed from healthy controls.

### Implication for future studies and clinical practice

While our study extends on previous results suggesting that increased relative abundance of *Eggerthella* and *Hungathella* as obtained by 16S rDNA sequencing plays a role in the pathophysiology of depression, further research is needed to unravel the underlying biological mechanisms. To characterize potential functional and metabolic shifts, deeper sequencing approaches such as shotgun metagenomic or metatranscriptomic sequencing will be essential.^9^ The gold standard to infer causal relationship is a randomized-controlled intervention. In rodent models, the transplantation of specific taxa for example through oral gavage - as performed elegantly with *Eggerthella* in a rodent model for rheumatoid arthritis by Balakrishnan et al. - can be utilized to investigate subsequent change in behavior, and systemic and central nervous changes in metabolites and inflammatory markers.^52^

Our findings have significant implications for clinical practice. The identification of specific gut microbiota as potential causal factors in MDD suggests that microbiome-targeted therapies, such as dietary interventions, probiotics, or fecal microbiota transplants, could offer novel treatment strategies at least in those share of patients where microbiome-induced inflammatory phenomena play a role. While these interventions are limited in their specificity, functional characterization of these taxa could identify systemic metabolites that could be targeted or replaced if depleted. Given the challenge of treatment-resistant depression, such microbiome-based therapies could provide significant benefits where traditional pharmacological approaches are insufficient.

### Limitations

The observational nature of the data, while enhanced by advanced causal inference techniques, still limits the ability to definitively establish causality. As outlined above, randomized controlled experiments are needed to confirm causality and explore the dynamic interactions between diet, microbiome, and mental health. Additionally, while our sample size is large and clinically well-characterized, further replication in diverse populations will enhance the generalizability of our findings, particularly since regional differences are known to contribute to impact gut-microbiome composition.

### Conclusions

In conclusion, our study provides robust evidence that specific gut microbiota, particularly *Eggerthella* and *Hungatella*, play an important role in the pathophysiology of MDD. These findings enhance our understanding of the biological underpinnings of depression and suggest potential avenues for innovative treatment approaches. Future research should focus on longitudinal and randomized controlled interventional studies to validate these causal relationships and develop effective microbiome-based therapies for depression. By integrating insights from the gut microbiome with existing psychiatric and metabolic research, we can move towards a more holistic understanding and treatment of MDD.

## Author Contributions

Conception and design: LFe, AHR, NRW, SET, DH, and TH

Acquisition of the data: NRW, LFi, SM, KF, TB, JG, EJL, FS, FTO, PU, LT, IN, RN, BS, NA, HJ, AJ, UD, and TK

Data preprocessing: LFe, MW, and CB

Data analysis: LFe (association / differential abundance analyses) and AHR (causal analyses)

Data interpretation: LFe, AHR, NRW, SET, MB, EN, DM, AAV, RN, AL, AR, SKS, JC, AF, BV, GC, AL, UD, TK, DH, and TH

Administrative, technical, or material support: CB, MB, EN, DM, AAV, RN, AL, and AF

Supervision: DH and TH

Funding acquisition: TK, UD, TH, and DH

Writing – original draft: LFe, AHR, NRW, SET, DH, and TH

Writing – review & editing: all authors

All authors had full access to all the data in the study and had final responsibility for the decision to submit for publication.

## Funding & Acknowledgements

This work was funded in part by the consortia grants from the German Research Foundation (DFG) SFB/TRR 393 (project grant no 521379614), the European Union’s Horizon 2020 Research and Innovation Program under grant agreement No. 847879 and the Hessian Ministry of Science and Arts (HMWK) LOEWE program (LOEWE Centre DYNAMIC).

This work is part of the German multicenter consortium “Neurobiology of Affective Disorders. A translational perspective on brain structure and function“, funded by the German Research Foundation (Deutsche Forschungsgemeinschaft DFG; Forschungsgruppe/Research Unit FOR2107).

Principal investigators (PIs) with respective areas of responsibility in the FOR2107 consortium are:

Work Package WP1, FOR2107/MACS cohort and brainimaging: Tilo Kircher (speaker FOR2107; DFG grant numbers KI588/14-1, KI588/14-2, KI588/20-1, KI588/22-1), Udo Dannlowski (co-speaker FOR2107; DA 1151/5-1, DA 1151/5-2, DA1151/6-1), Axel Krug (KR 3822/5-1, KR 3822/7-2), Igor Nenadic (NE2254/1-2, NE2254/3-1, NE2254/4-1), Carsten Konrad (KO 4291/3-1). WP2, animal phenotyping: Markus Wöhr (WO 1732/4-1, WO 1732/4-2), Rainer Schwarting (SCHW 559/14-1, SCHW 559/14-2). WP3, miRNA: Gerhard Schratt (SCHR 1136/3-1, 1136/3-2). WP4, immunology, mitochondriae: Judith Alferink (AL 1145/5-2), Carsten Culmsee (CU 43/9-1, CU 43/9-2), Holger Garn (GA 545/5-1, GA 545/7-2). WP5, genetics: Marcella Rietschel (RI 908/11-1, RI 908/11-2), Markus Nöthen (NO 246/10-1, NO 246/10-2), Stephanie Witt (WI 3439/3-1, WI 3439/3-2). WP6, multi-method data analytics: Andreas Jansen (JA 1890/7-1, JA 1890/7-2), Tim Hahn (HA 7070/2-2), Bertram Müller-Myhsok (MU1315/8-2), Astrid Dempfle (DE 1614/3-1, DE 1614/3-2). CP1, biobank: Petra Pfefferle (PF 784/1-1, PF 784/1-2), Harald Renz (RE 737/20-1, 737/20-2). CP2, administration. Tilo Kircher (KI 588/15-1, KI 588/17-1), Udo Dannlowski (DA 1151/6-1), Carsten Konrad (KO 4291/4-1).

Data access and responsibility: All PIs take responsibility for the integrity of the respective study data and their components. All authors and coauthors had full access to all study data.

Acknowledgements and members by Work Package (WP):

WP1: Henrike Bröhl, Katharina Brosch, Bruno Dietsche, Rozbeh Elahi, Jennifer Engelen, Ulrika Evermann, Sabine Fischer, Jessica Heinen, Svenja Klingel, Felicitas Meier, Tina Meller, Julia-Katharina Pfarr, Kai Ringwald, Torsten Sauder, Simon Schmitt, Frederike Stein, Lea Teutenberg, Florian Thomas-Odenthal, Annette Tittmar, Adrian Wroblewski, Paula Usemann, Dilara Yüksel (Dept. of Psychiatry, Marburg University). Mechthild Wallnig, Rita Werner (Core-Facility Brainimaging, Marburg University). Carmen Schade-Brittinger, Maik Hahmann (Coordinating Centre for Clinical Trials, Marburg). Michael Putzke (Psychiatric Hospital, Friedberg). Rolf Speier, Lutz Lenhard (Psychiatric Hospital, Haina). Birgit Köhnlein (Psychiatric Practice, Marburg). Peter Wulf, Jürgen Kleebach, Achim Becker (Psychiatric Hospital Hephata, Schwalmstadt-Treysa). Ruth Bär (Care facility Bischoff, Neukirchen). Matthias Müller, Michael Franz, Siegfried Scharmann, Anja Haag, Kristina Spenner, Ulrich Ohlenschläger (Psychiatric Hospital Vitos, Marburg). Matthias Müller, Michael Franz, Bernd Kundermann (Psychiatric Hospital Vitos, Gießen). Christian Bürger, Katharina Dohm, Fanni Dzvonyar, Verena Enneking, Stella Fingas, Kira Flinkenflügel, Katharina Förster, Janik Goltermann, Dominik Grotegerd, Hannah Lemke, Susanne Meinert, Nils Opel, Ronny Redlich, Jonathan Repple, Katharina Thiel, Kordula Vorspohl, Bettina Walden, Lena Waltemate, Alexandra Winter, Dario Zaremba (Dept. of Psychiatry, University of Münster). Harald Kugel, Jochen Bauer, Walter Heindel, Birgit Vahrenkamp (Dept. of Clinical Radiology, University of Münster). Gereon Heuft, Gudrun Schneider (Dept. of Psychosomatics and Psychotherapy, University of Münster). Thomas Reker (LWL-Hospital Münster). Gisela Bartling (IPP Münster). Ulrike Buhlmann (Dept. of Clinical Psychology, University of Münster), Robert Nitsch (Institute for Translational Neuroscience, University of Münster).

WP2: Marco Bartz, Miriam Becker, Christine Blöcher, Annuska Berz, Moria Braun, Ingmar Conell, Debora dalla Vecchia, Darius Dietrich, Ezgi Esen, Sophia Estel, Jens Hensen, Ruhkshona Kayumova, Theresa Kisko, Rebekka Obermeier, Anika Pützer, Nivethini Sangarapillai, Özge Sungur, Clara Raithel, Tobias Redecker, Vanessa Sandermann, Finnja Schramm, Linda Tempel, Natalie Vermehren, Jakob Vörckel, Stephan Weingarten, Maria Willadsen, Cüneyt Yildiz (Faculty of Psychology, Marburg University).

WP4: Jana Freff (Dept. of Psychiatry, University of Münster). Susanne Michels, Goutham Ganjam, Katharina Elsässer (Faculty of Pharmacy, Marburg University). Felix Ruben Picard, Nicole Löwer, Thomas Ruppersberg (Institute of Laboratory Medicine and Pathobiochemistry, Marburg University).

WP5: Helene Dukal, Christine Hohmeyer, Lennard Stütz, Viola Lahr, Fabian Streit, Josef Frank, Lea Sirignano (Dept. of Genetic Epidemiology, Central Institute of Mental Health, Medical Faculty Mannheim, Heidelberg University). Stefanie Heilmann-Heimbach, Stefan Herms, Per Hoffmann (Institute of Human Genetics, University of Bonn, School of Medicine & University Hospital Bonn). Andreas J. Forstner (Institute of Human Genetics, University of Bonn, School of Medicine & University Hospital Bonn; Centre for Human Genetics, Marburg University).

WP6: Anastasia Benedyk, Miriam Bopp, Roman Keßler, Maximilian Lückel, Verena Schuster, Christoph Vogelbacher (Dept. of Psychiatry, Marburg University). Jens Sommer, Olaf Steinsträter (Core-Facility Brainimaging, Marburg University). Thomas W.D. Möbius (Institute of Medical Informatics and Statistics, Kiel University).

CP1: Julian Glandorf, Fabian Kormann, Arif Alkan, Fatana Wedi, Lea Henning, Alena Renker, Karina Schneider, Elisabeth Folwarczny, Dana Stenzel, Kai Wenk, Felix Picard, Alexandra Fischer, Sandra Blumenau, Beate Kleb, Doris Finholdt, Elisabeth Kinder, Tamara Wüst, Elvira Przypadlo, Corinna Brehm (Comprehensive Biomaterial Bank Marburg, Marburg University).

The FOR2107 cohort project (WP1) was approved by the Ethics Committees of the Medical Faculties, University of Marburg (AZ: 07/14) and University of Münster (AZ: 2014-422-b-S).

SET was funded by the Leistungszentrum Innovative Therapeutics (TheraNova) funded by the Fraunhofer Society and the Hessian Ministry of Science and Art, the Bundesministerium für Bildung und Forschung (BMBF, Federal Ministry of Education) - 01EO2102 INITIALISE Advanced Clinician Scientist Program and the REISS foundation.

RN was funded by the German Research Foundation (DFG/CRC 1451/A07).

Biomedical financial interests or potential conflicts of interest: Tilo Kircher received unrestricted educational grants from Servier, Janssen, Recordati, Aristo, Otsuka, neuraxpharm. Markus Wöhr is scientific advisor of Avisoft Bioacoustics. Sarah Kittel-Schneider received speaker’s honoraria from Janssen. Andreas Reif has received honoraria for lectures and/or advisory boards from Janssen, Boehringer Ingelheim, COMPASS, SAGE/Biogen, LivaNova, Medice, Shire/Takeda, MSD and cyclerion. Also, he has received research grants from Medice and Janssen.

Microbiota sequencing @IKMB received infrastructure support from the DFG Research Unit 5042 „miTarget" and the DFG Excellence Cluster 2167 "Precision Medicine in Chronic Inflammation" (PMI).

## Data Availability

All data produced in the present study are available upon reasonable request to the authors

## Supplementary Material

### Supplementary Methods

#### sMethods 1. Study information and sample exclusion criteria

The ethics committees of the medical faculties of the University of Marburg, Germany, and the University of Münster, Germany, approved the study. Participants received financial compensation and gave written and informed consent. Patients were recruited from local in- and outpatient services and either fulfilled the DSM-IV criteria for an acute major depressive episode or had a lifetime history of a major depressive episode. Participants with a history of neurological (e.g., concussion, stroke, tumor, neuro-inflammatory diseases) or medical conditions (e.g., cancer, chronic inflammatory or autoimmune diseases, heart diseases, diabetes mellitus, infections), as well as those who self-identified as non-Caucasian, were excluded from the analysis. Non-Caucasian participants were excluded since the FOR2107 MACS cohort was originally focused on genetic and neuroimaging analyses, thus providing greater genetic homogeneity. The exclusion criteria were the same for both the healthy and depressive participants. Additionally, healthy participants were further excluded if they had a current or past history of psychiatric illness. A sample of 1,801 participants remained for the analyses.

#### sMethods 2. Causal Discovery Analysis

The FCI algorithm starts with the adjacency phase, conducting multiple conditional independence tests to infer the *skeleton* of the underlying causal model – an undirected graph encoding the conditional independencies observed in the data. Initially, a complete graph is created, where vertices represent observed variables, and each distinct pair of vertices is connected by an edge. Then, considering each pair {V_i, V_j} of observed variables, the algorithm traverses through all potential subsets of other observed variables aiming to identify a set S_ij such that V_i becomes conditionally independent of V_j given S_ij. If such a subset exists, the edge between V_i and V_j is removed. In our analysis, we used the stable version of the algorithm, which produces a fully order-independent final skeleton, as shown by Colombo and Maathuis^64^.

As our dataset includes both binary and continuous variables, we use the symmetric conditional independence test for mixed data proposed by Tsagris et al.^44^, available in the MXM R package^45^. This test evaluates the conditional independence of two variables, V_i and V_j, given a set of variables S_ij, by testing two null hypotheses: H0_1: P(V_i| S_ij) = P(V_i|V_j, S_ij) and H0_2: P(V_j| S_ij) = P(V_j|V_i, S_ij). The null hypothesis H0_1 is tested using a nested likelihood-ratio test comparing a reduced model (where V_i is regressed on S_ij) against a full model (where V_i is regressed on both S_ij and V_j). Similarly, H0_2 is tested by reversing the roles of V_i and V_j. In general, the p-values p1 and p2 from the tests for H0_1 and H0_2, respectively, tend to be identical only asymptotically. To correct any asymmetry in limited data scenarios, we follow the authors’ technique of merging dependent p-values. Such method calculates the combined p-value as min(2 * min(p1, p2), max(p1, p2)) and has demonstrated superior learning accuracy when compared to alternative methods.

Tsagris et al.’s^44^ test relies on linear or generalized linear functions, depending on the nature of the outcome variable. For instance, a test involving a binary outcome relies on a logistic regression, while a test involving a continuous outcome relies on a Gaussian linear regression. To address deviations from normality when BMI is the outcome, we transformed it using a rank-based inverse normal transformation. For cases where a microbial taxa acts as an outcome variable, we implemented two additional likelihood-ratio tests, one based on LinDA and another based on ZicoSeq. Importantly, to ensure thorough adjustment for the influence of sex, age, and library size, even when minor, we enforce their inclusion in the conditioning set for all conditional independence tests. Age and sex stand as unique covariates, as they are known for not being caused by any other variables, and therefore, conditioning on them can never introduce biases such as collider bias. Additionally, library size reflects inherent artifacts of the sequencing platform, making it crucial to adjust for it to correct potential compositional biases.

After constructing the underlying model’s skeleton, the FCI algorithm advances to the orientation phase, where a set of 10 orientation rules^20,41^ are iteratively applied until no further edge marks can be identified. In general, however, not all edge marks can be established due to the existence of multiple graphical models that can entail the same set of conditional independencies. This results in a class of statistically equivalent models termed Markov equivalent class (MEC). As a result, FCI constructs a Partial Ancestral Graph (PAG) representing all ancestral (causal) and non-ancestral relationships common to models within the most plausible MEC. In a PAG, arrowheads represent definite non-ancestral (non-causal) relationships, while tails represent definite ancestral (causal) relationships. A circle ("o") denotes non-invariant edge marks, indicating the existence within the MEC of both a model where the edge mark is a tail and another model where the same edge mark is an arrowhead.

A PAG represents a class of the most probable models based on the available data. Remarkably, all models in this class fit the data equally well, as they entail the same set of observed conditional independencies, rendering them statistically indistinguishable. Whenever ancestral and non-ancestral relationships are shared across all models within this equivalence class, they are represented as non-circle edges in the PAG.

The process of learning a PAG through causal discovery algorithms is intricate. Most algorithms, including the FCI, rely on an assumption known as *faithfulness* to ensure that the inferred PAG accurately represents the MEC of the true underlying causal model. Such an assumption asserts that the set of conditional independence relations inferred during the learning process is truly satisfied by the underlying model. This premise, however, is often violated in real-world scenarios, especially when dealing with datasets of finite sample size, where statistical tests may lack the power to draw accurate conclusions. Misidentified independencies or those established under an inaccurate conditioning set can lead not only to potentially multiple wrong edge orientations^65,66^, but also to violations of the inherent PAG properties and inconsistencies in the set of implied conditional independencies through m-separation^22^. Remarkably, this issue greatly intensifies when dealing with larger graphs due to both the higher number of conditional independence tests and the substantial decrease in statistical power as the conditioning set size grows.

To ensure the reliability of our analysis, we employ different approaches. First, we evaluate marginal causal consistency following the approach by Roumpelaki et al.^46^. Specifically, we apply the FCI algorithm considering all possible subsets of the selected variables and verify the degree of agreement of causal relations among the all marginals. As FCI’s soundness holds despite latent confounding, if there is no statistical error, then every definite ancestral (causal) or non-ancestral (non-causal) relationship established in a PAG over a particular subset of variables should hold true in the full underlying model and must consistently align with any other definite relationships established across different subsets of variables. Consequently, if conflicting edge orientations arise, conclusions should be approached cautiously as they might solely stem from statistical inaccuracies. To further enhance reliability, we also implemented extensive tests to verify whether each inferred PAG aligns with its characterization of a class of Maximal Ancestral Graphs^20^ and only implies conditional independencies that are consistent with the observed data. Any instances revealing violations or inconsistencies are excluded from our analysis. Finally, we compare all PAGs that passed our validity and robustness tests with the ones obtained by the conservative variant of the FCI algorithm^47^, which performs additional tests to identify ambiguous triplets and then selectively applies the orientation rules only when supported by unambiguous triplets.

#### sMethods 3. Causal Effect Identification and Estimation

As previously discussed, a PAG provides a qualitative description of the ancestral and non-ancestral relationships among variables, common across all models consistent with observed data. To quantitatively analyze causal effects, it is crucial to assess their identifiability. Specifically, a causal effect can be inferred from a PAG if it is uniquely computable within its equivalence class using expressions based solely on observational (conditional) probabilities.

Sound and complete tools have been developed for causal effect identification from PAGs. These include the generalized backdoor criterion^48^, adjustment criterion^21^, as well as causal calculus and (conditional) effect identification algorithms^22^.

The generalized backdoor criterion helps us determine the causal effect of an exposure variable X on an outcome variable Y from a PAG P. It searches for a set Z of variables that are not descendants of X and that block (in the sense of d-separation) all potential confounding paths between X and Y in P. If this set Z is identified, we can compute the causal effect of X on Y through an adjustment formula that incorporates Z. When analyzing the causal effect of X on Y conditioned on another set S, the adjustment set Z must include S. For further details, refer to Pearl^19^. The estimation of these causal effects is facilitated by the marginal effects R package (Arel-Bundock et al.), typically modeling relationships among variables with (generalized) linear regression models and computing average estimates through Monte Carlo methods.

#### sMethods 4. Summary table of psychopharmacological medication. Participants taking any of these medications were considered “medicated”

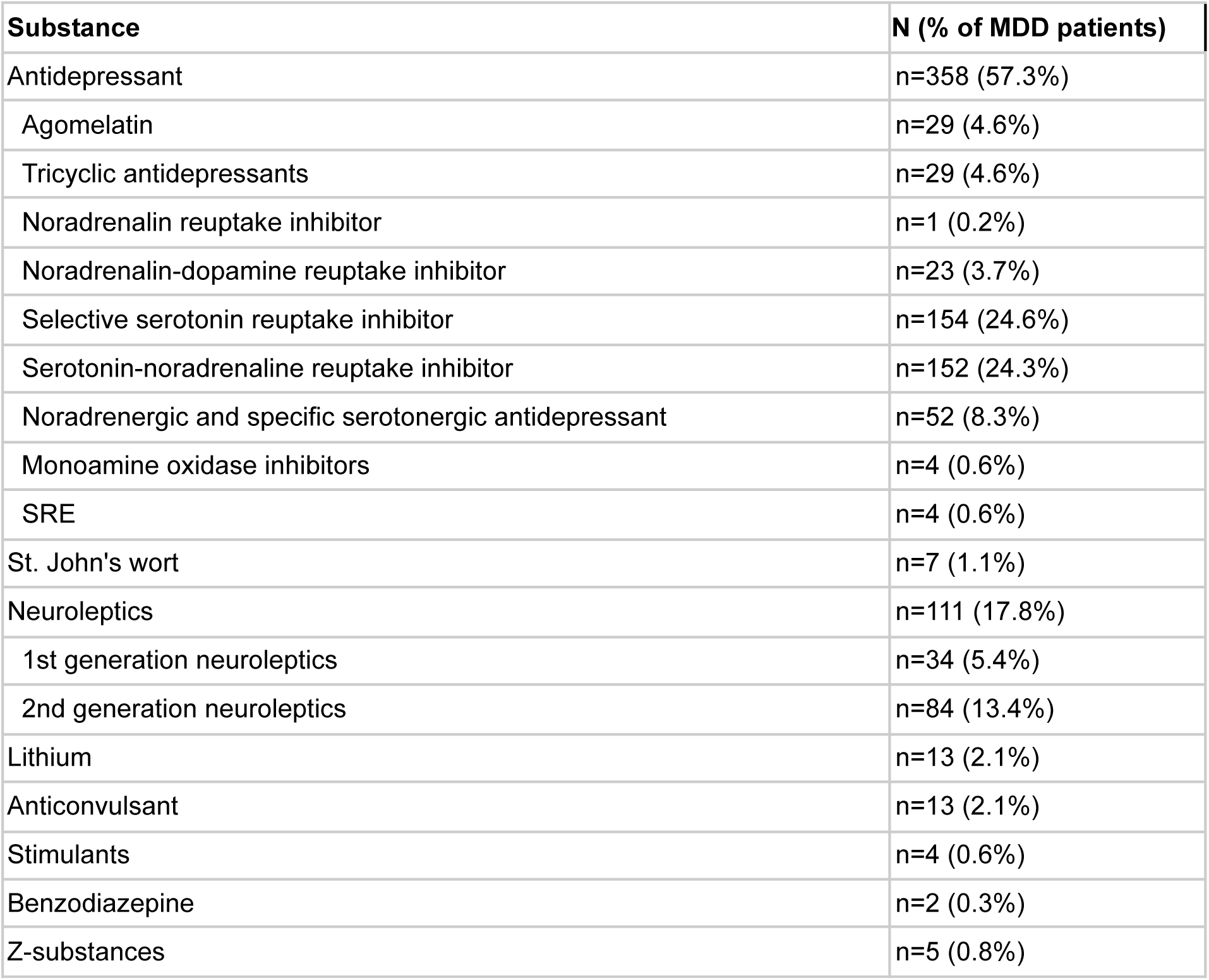

| **Substance** | **N (% of MDD patients)** |
| --- | --- |
| Antidepressant | n=358 (57.3%) |
| Agomelatin | n=29 (4.6%) |
| Tricyclic antidepressants | n=29 (4.6%) |
| Noradrenalin reuptake inhibitor | n=1 (0.2%) |
| Noradrenalin-dopamine reuptake inhibitor | n=23 (3.7%) |
| Selective serotonin reuptake inhibitor | n=154 (24.6%) |
| Serotonin-noradrenaline reuptake inhibitor | n=152 (24.3%) |
| Noradrenergic and specific serotonergic antidepressant | n=52 (8.3%) |
| Monoamine oxidase inhibitors | n=4 (0.6%) |
| SRE | n=4 (0.6%) |
| St. John's wort | n=7 (1.1%) |
| Neuroleptics | n=111 (17.8%) |
| 1st generation neuroleptics | n=34 (5.4%) |
| 2nd generation neuroleptics | n=84 (13.4%) |
| Lithium | n=13 (2.1%) |
| Anticonvulsant | n=13 (2.1%) |
| Stimulants | n=4 (0.6%) |
| Benzodiazepine | n=2 (0.3%) |
| Z-substances | n=5 (0.8%) |

### Supplementary Results

**sTable 1:** Results of the logistic regression for descriptive statistics.

Supplementary Results **sTable 1:** Results of the logistic regression for descriptive statistics
|  | Estimate | Std. Error | z value | Pr(>\|z\|) |
| --- | --- | --- | --- | --- |
| (Intercept) | -2.792 | 0.688 | -4.056 | 4.99e-05 |
| Shannon Index | -0.165 | 0.179 | -0.921 | 0.357 |
| Age | 0.86e-02 | 0.48e -02 | 1.794 | 0.073 |
| Sex | 0.041 | 0.122 | 0.333 | 0.739 |
| BMI | 0.068 | 0.0121 | 5.664 | 1.48e-08 |
| Library Size | 0.39e-05 | 0.72e-05 | 0.539 | 0.590 |
| Site | 0.638 | 0.131 | 4.863 | 1.16e-06 |

**sFigure 1:**
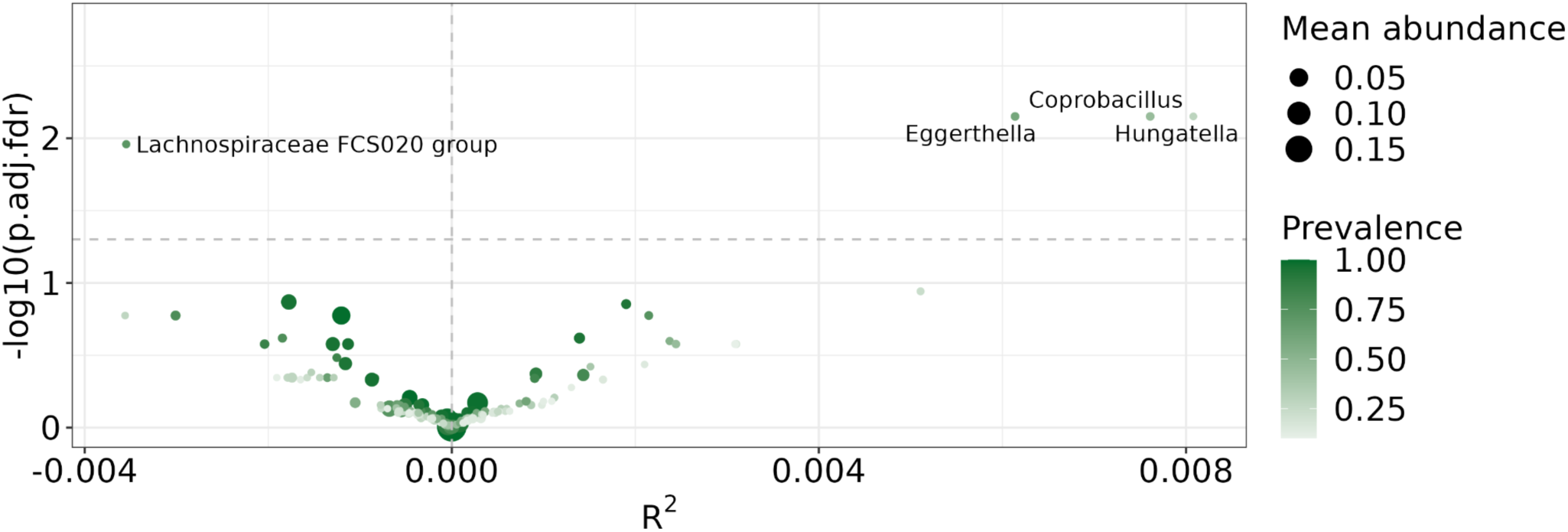
Results of the ZicoSeq abundance analysis. Adjusted p-values given by ZicoSeq for each genus with color intensity indicating prevalence and bubble size mean abundance. Significant (p<0.05) genera lie above the horizontal dashed line and are named. R² values indicate whether the genus was more abundant in the MDD group (right of the vertical dashed line) or in the HC group (left of the vertical dashed line). Eggerthella mean relative abundance in healthy controls was ∼0.00054 with a prevalence of ∼0.56832, while in the MDD group the mean relatiive abundance was ∼0.00087 with a prevalence of 0.63040; Hungatella mean relative abundance in healthy controls was ∼0.00006 with a prevalence of ∼0.24068, while in the MDD group the mean relatiive abundance was ∼0.00012 with a prevalence of 0.33600

**sFigure 2:**
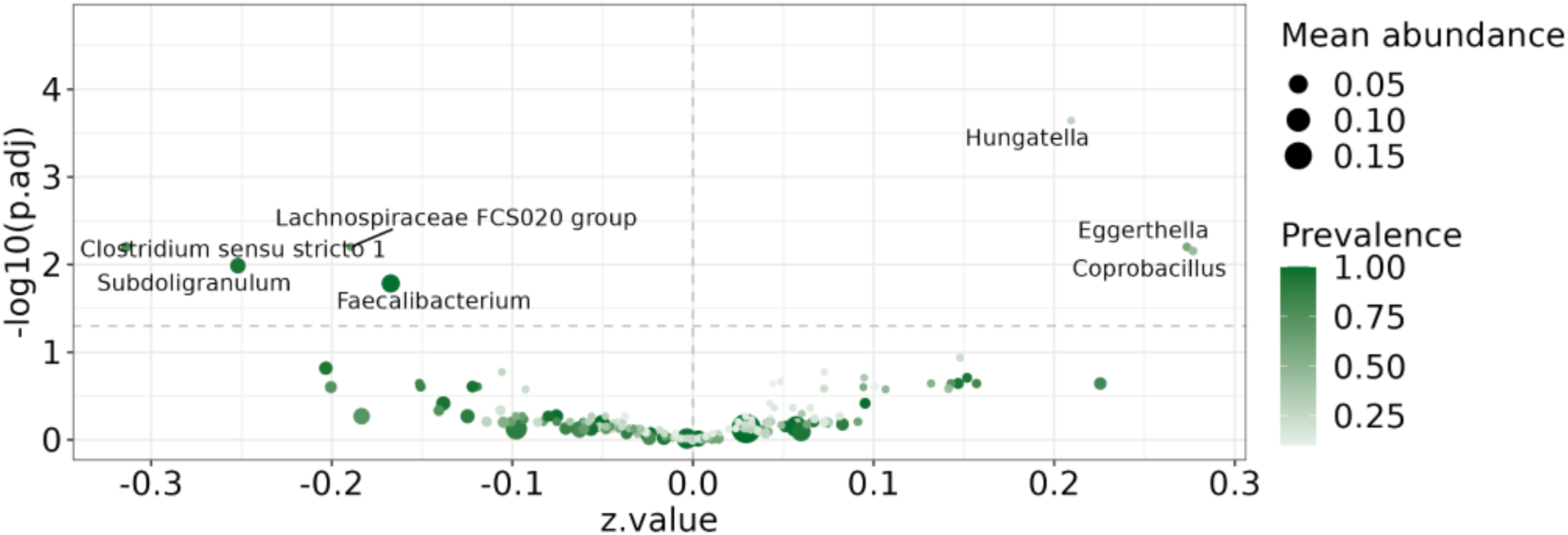
Results of the LinDA analysis. Adjusted p-values given by LinDA for each genus with color intensity indicating prevalence and bubble size mean abundance. Significant (p<0.05) genera lie above the horizontal dashed line and are named. Z-values indicate whether the genus was more abundant in the MDD group (right of the vertical dashed line) or in the HC group (left of the vertical dashed line).

**sTable 2:** Results of medication analysis of *Eggerthella* and *Hungatella.* The data was divided into three groups, the medicated group (N=360), the unmedicated group (N=244) and the unaffected controls (N=626). Only participants with complete information on age, sex, BMI, and medication index were included in this analysis. The group mentioned first on the comparison column was the reference group. A positive estimate indicates higher abundance in the non-reference group while a negative estimate indicates higher abundance in the reference group. The estimates and the respective confidence intervals were obtained from logistic regression models, while q-values were obtained through FDR correction. The models were adjusted for age, sex, BMI and collection site.

| Genus | Comparison | Mean clr abundance group 1 | Mean clr abundance group 2 | Estimate | 95% CI |  | p value | q value |
| --- | --- | --- | --- | --- | --- | --- | --- | --- |
| *Eggerthella* | Unaffected – Unmedicated | -0.413 | -0.013 | 0.168 | 0.020 | 0.316 | 0.026 | 0.040 |
|  | Unaffected – Medicated | -0.413 | 0.032 | 0.243 | 0.108 | 0.378 | < 0.001 | 0.001 |
|  | Unmedicated – medicated | -0.013 | 0.032 | 0.110 | -0.061 | 0.280 | 0.207 | 0.248 |
| *Hungatella* | Unaffected Unmedicated | -1.190 | -0.986 | 0.238 | 0.094 | 0.382 | 0.001 | 0.002 |
|  | Unaffected – Medicated | -1.190 | -0.911 | 0.322 | 0.183 | 0.460 | < 0.001 | < 0.001 |
|  | Unmedicated – medicated | -0.986 | -0.911 | 0.094 | -0.077 | 0.265 | 0.282 | 0.282 |

**sTable 3:**
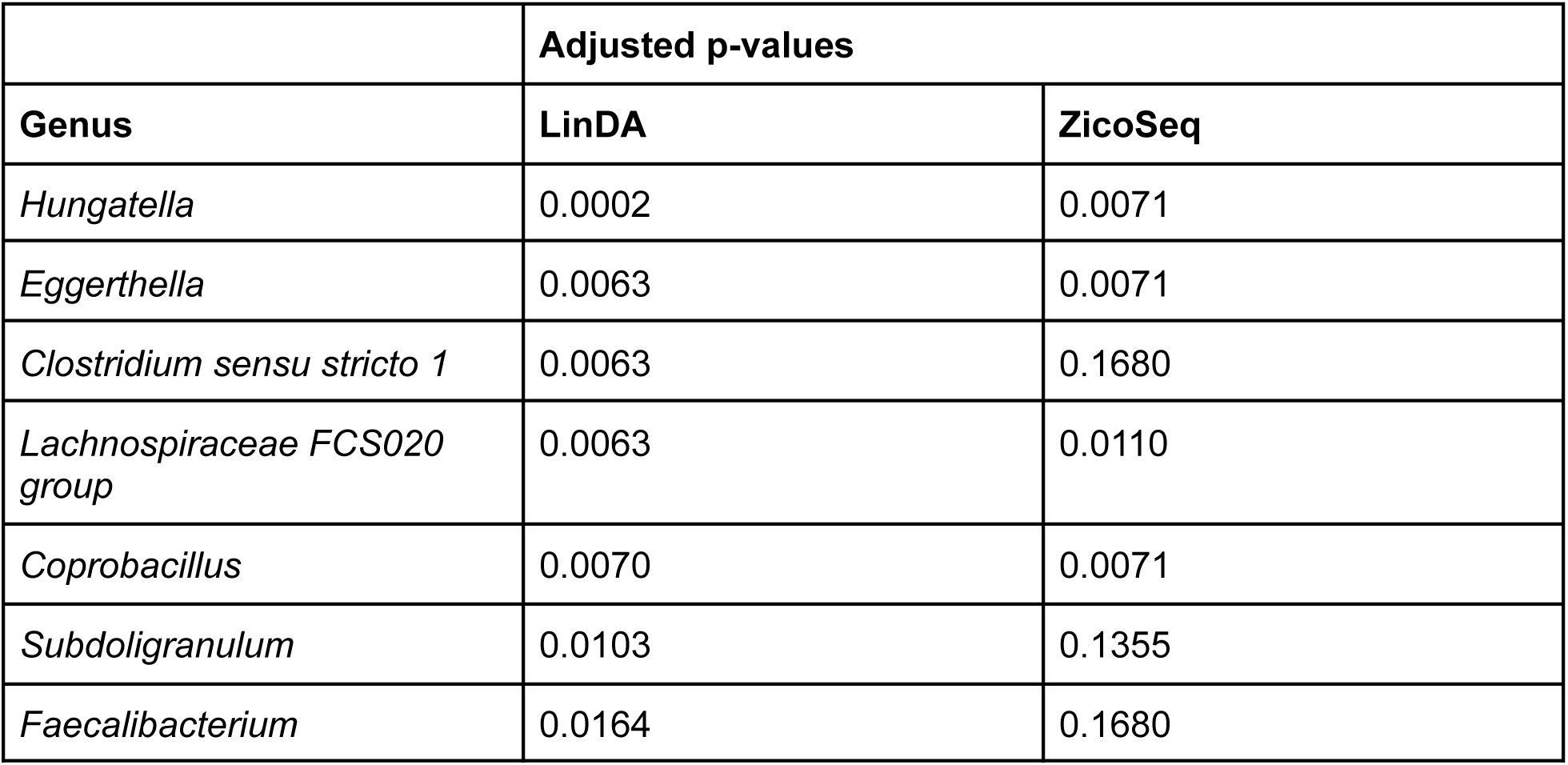
Results of the Differential Abundance Analysis using the LiNDA and ZicoSeq tools.

**sFigure 3:**
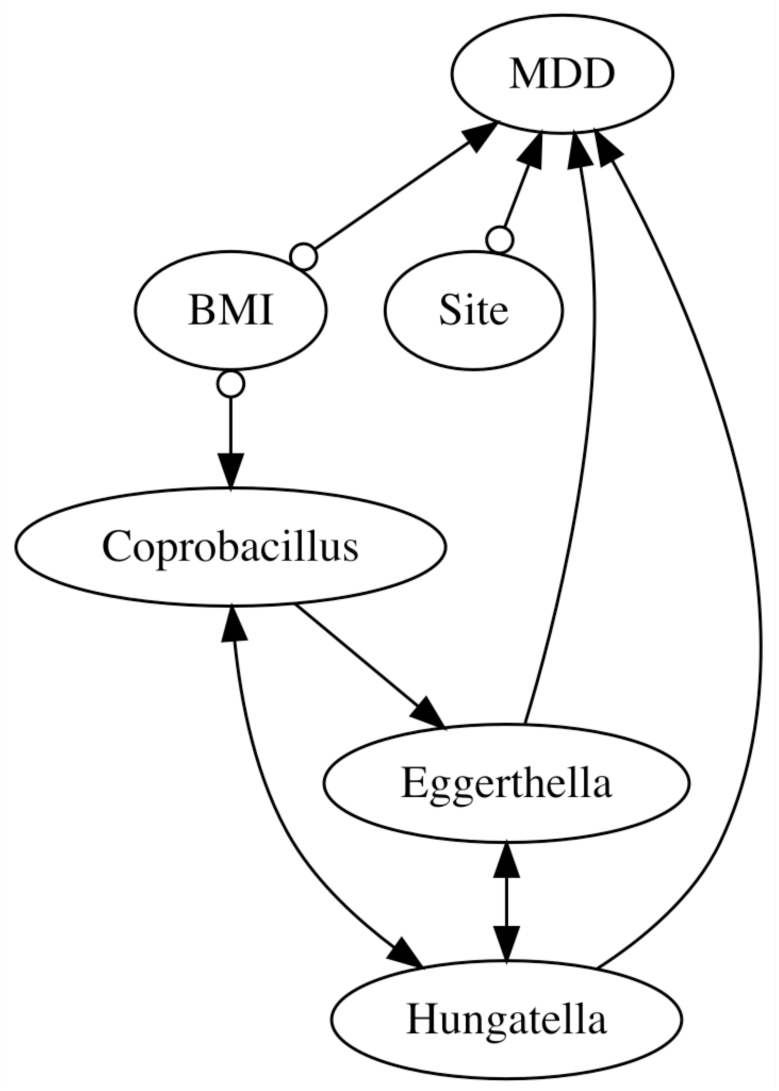
Conservative FCI’s resulting PAG. Conservative FCI’s resulting PAG over MDD, BMI, Site and all microbial taxa robustly identified as causes of MDD, namely *Eggertella*, *Hungatella*, and *Copobracillus*. Conditional independence tests for taxa variables rely on LinDA and consistently include age, sex, and library size as part of the conditioning set.

**sTable4:** Relationships from PAGs that successfully passed our validity and robustness tests, and were identically inferred by both FCI and its conservative variant, while using a conditional independence test for microbial taxa variables based on <u>LinDA</u>. The column "0" indicates no edge. The column "sum" indicates the total number of PAGs that passed our tests, out of the 31 constructed ones.

| relation | 0 | o-o | <-o | -o | o-> | <-> | -> | o- | <- | - | sum |
| --- | --- | --- | --- | --- | --- | --- | --- | --- | --- | --- | --- |
| MDD,BMI | 0 | 7 | 20 | 0 | 0 | 0 | 0 | 0 | 0 | 0 | 27 |
| MDD,Site | 0 | 9 | 18 | 0 | 0 | 0 | 0 | 0 | 0 | 0 | 27 |
| BMI,Site | 29 | 0 | 0 | 0 | 0 | 0 | 0 | 0 | 0 | 0 | 29 |
| MDD,Lachnospiraceae_FCS020_group | 0 | 12 | 12 | 0 | 0 | 0 | 0 | 0 | 0 | 0 | 24 |
| BMI,Lachnospiraceae_FCS020_group | 27 | 0 | 0 | 0 | 0 | 0 | 0 | 0 | 0 | 0 | 27 |
| MDD,Coprobacillus | 2 | 18 | 4 | 0 | 0 | 0 | 0 | 0 | 0 | 0 | 24 |
| BMI,Coprobacillus | 0 | 7 | 0 | 0 | 20 | 0 | 0 | 0 | 0 | 0 | 27 |
| MDD,Eggerthella | 0 | 22 | 0 | 0 | 0 | 0 | 0 | 0 | 2 | 0 | 24 |
| BMI,Eggerthella | 12 | 15 | 0 | 0 | 0 | 0 | 0 | 0 | 0 | 0 | 27 |
| MDD,Hungatella | 0 | 25 | 0 | 0 | 0 | 0 | 0 | 0 | 2 | 0 | 27 |
| BMI,Hungatella | 29 | 0 | 0 | 0 | 0 | 0 | 0 | 0 | 0 | 0 | 29 |
| Site,Lachnospiraceae_FCS020_group | 27 | 0 | 0 | 0 | 0 | 0 | 0 | 0 | 0 | 0 | 27 |
| Site,Coprobacillus | 27 | 0 | 0 | 0 | 0 | 0 | 0 | 0 | 0 | 0 | 27 |
| Site,Eggerthella | 27 | 0 | 0 | 0 | 0 | 0 | 0 | 0 | 0 | 0 | 27 |
| Site,Hungatella | 29 | 0 | 0 | 0 | 0 | 0 | 0 | 0 | 0 | 0 | 29 |
| Lachnospiraceae_FCS020_group,Coprobacillus | 0 | 14 | 0 | 0 | 8 | 2 | 0 | 0 | 0 | 0 | 24 |
| Lachnospiraceae_FCS020_group,Eggerthella | 1 | 21 | 0 | 0 | 0 | 2 | 0 | 0 | 0 | 0 | 24 |
| Lachnospiraceae_FCS020_group,Hungatella | 0 | 27 | 0 | 0 | 0 | 0 | 0 | 0 | 0 | 0 | 27 |
| Coprobacillus,Eggerthella | 0 | 16 | 0 | 0 | 0 | 0 | 8 | 0 | 0 | 0 | 24 |
| Coprobacillus,Hungatella | 0 | 13 | 8 | 0 | 0 | 6 | 0 | 0 | 0 | 0 | 27 |
| Eggerthella,Hungatella | 0 | 21 | 0 | 0 | 0 | 6 | 0 | 0 | 0 | 0 | 27 |

**sTable 5:** Relationships from PAGs that successfully passed our validity and robustness tests, and were identically inferred by both FCI and its conservative variant, while using a conditional independence test for microbial taxa variables based on <u>ZicoSeq</u>. The column "0" indicates no edge. The column "sum" indicates the total number of PAGs that passed our tests, out of the 31 constructed ones.

| relation | 0 | o-o | <-o | -o | o-> | <-> | -> | o- | <- | - | sum |
| --- | --- | --- | --- | --- | --- | --- | --- | --- | --- | --- | --- |
| MDD,BMI | 0 | 6 | 11 | 0 | 0 | 0 | 0 | 0 | 0 | 0 | 17 |
| MDD,Site | 0 | 10 | 10 | 0 | 0 | 0 | 0 | 0 | 0 | 0 | 20 |
| BMI,Site | 21 | 0 | 0 | 0 | 0 | 0 | 0 | 0 | 0 | 0 | 21 |
| MDD,Lachnospiraceae_FCS020_group | 0 | 8 | 11 | 0 | 0 | 0 | 0 | 0 | 0 | 0 | 19 |
| BMI,Lachnospiraceae_FCS020_group | 20 | 0 | 0 | 0 | 0 | 0 | 0 | 0 | 0 | 0 | 20 |
| MDD,Coprobacillus | 0 | 16 | 4 | 0 | 0 | 0 | 0 | 0 | 0 | 0 | 20 |
| BMI,Coprobacillus | 0 | 12 | 0 | 0 | 9 | 0 | 0 | 0 | 0 | 0 | 21 |
| MDD,Eggerthella | 0 | 12 | 4 | 0 | 0 | 0 | 0 | 0 | 0 | 0 | 16 |
| BMI,Eggerthella | 3 | 13 | 0 | 0 | 0 | 0 | 0 | 0 | 0 | 0 | 16 |
| MDD,Hungatella | 0 | 20 | 0 | 0 | 0 | 0 | 0 | 0 | 0 | 0 | 20 |
| BMI,Hungatella | 21 | 0 | 0 | 0 | 0 | 0 | 0 | 0 | 0 | 0 | 21 |
| Site,Lachnospiraceae_FCS020_group | 22 | 0 | 0 | 0 | 0 | 0 | 0 | 0 | 0 | 0 | 22 |
| Site,Coprobacillus | 22 | 0 | 0 | 0 | 0 | 0 | 0 | 0 | 0 | 0 | 22 |
| Site,Eggerthella | 20 | 0 | 0 | 0 | 0 | 0 | 0 | 0 | 0 | 0 | 20 |
| Site,Hungatella | 22 | 0 | 0 | 0 | 0 | 0 | 0 | 0 | 0 | 0 | 22 |
| Lachnospiraceae_FCS020_group,Coprobacillus | 0 | 10 | 0 | 0 | 9 | 0 | 0 | 0 | 0 | 0 | 19 |
| Lachnospiraceae_FCS020_group,Eggerthella | 17 | 0 | 0 | 0 | 0 | 0 | 0 | 0 | 0 | 0 | 17 |
| Lachnospiraceae_FCS020_group,Hungatella | 0 | 13 | 0 | 0 | 6 | 0 | 0 | 0 | 0 | 0 | 19 |
| Coprobacillus,Eggerthella | 0 | 18 | 0 | 0 | 0 | 0 | 0 | 0 | 0 | 0 | 18 |
| Coprobacillus,Hungatella | 0 | 19 | 0 | 0 | 0 | 0 | 0 | 0 | 0 | 0 | 19 |
| Eggerthella,Hungatella | 6 | 6 | 0 | 0 | 6 | 0 | 0 | 0 | 0 | 0 | 18 |

#### Effects of Continuous Abundances of Eggerthella and Hungatella on MDD

The estimated probabilities of experiencing MDD given a specific level "e" of *Eggerthella* abundance set by an intervention, expressed as P(MDD=1|do(*Eggerthela*=e)), and given a specific level "h" of *Hungatella* abundance set by an intervention, expressed as P(MDD=1|do(*Hungatella*=h)), were based on a logistic regression of MDD on *Eggerthella* and *Hungatella* abundances, and BMI. See *Supplementary Table 6* for the coefficient estimates and respective standard errors and p-values. Notably, all coefficients are statistically significant at the 5% level. It is important to emphasize that Age, Sex, and Library Size, when included in the model, do not reach statistical significance at the 5% threshold. In this model, the odds of MDD increases by approximately 1.008 with a one unit increase in the *Eggerthella* abundance, 1.056 with a one unit increase in *Hungatella* abundance, and 1.068 with a one unit increase in BMI.

The estimation of P(MDD=1|do(*Eggerthela*=e), catBMI=c) is based on a logistic regression of MDD on BMI Category as well as *Eggerthella* and *Hungatella* abundances. The coefficient estimates and their respective p-values are detailed in *Supplementary Table 7*. In this model, the odds of MDD still increases by approximately 1.008 with a one unit increase in the *Eggerthella* abundance, and 1.056 with a one unit increase in *Hungatella* abundance. Moreover, the odds of MDD increase by approximately 1.296 when moving from Non-Obese to Overweight and by approximately 2.596 when moving from Overweight to Obese.

**sTable 6:** Coefficient estimates and their respective p-values of a logistic regression of MDD on *Eggerthella* and *Hungatella* abundances, and BMI.

|  | Estimate | Std. Error | z value | Pr(>\|z\|) |
| --- | --- | --- | --- | --- |
| **(Intercept)** | -1.850900 | 0.296310 | -6.2464 | 4.2000e-10 |
| **BMI** | 0.007859 | 0.002529 | 3.1070 | 1.8898e-03 |
| **Eggerthella** | 0.065609 | 0.011529 | 5.6908 | 1.2647e-08 |
| **Hungatella** | 0.055294 | 0.015237 | 3.6289 | 2.8462e-04 |

**sTable 7:**
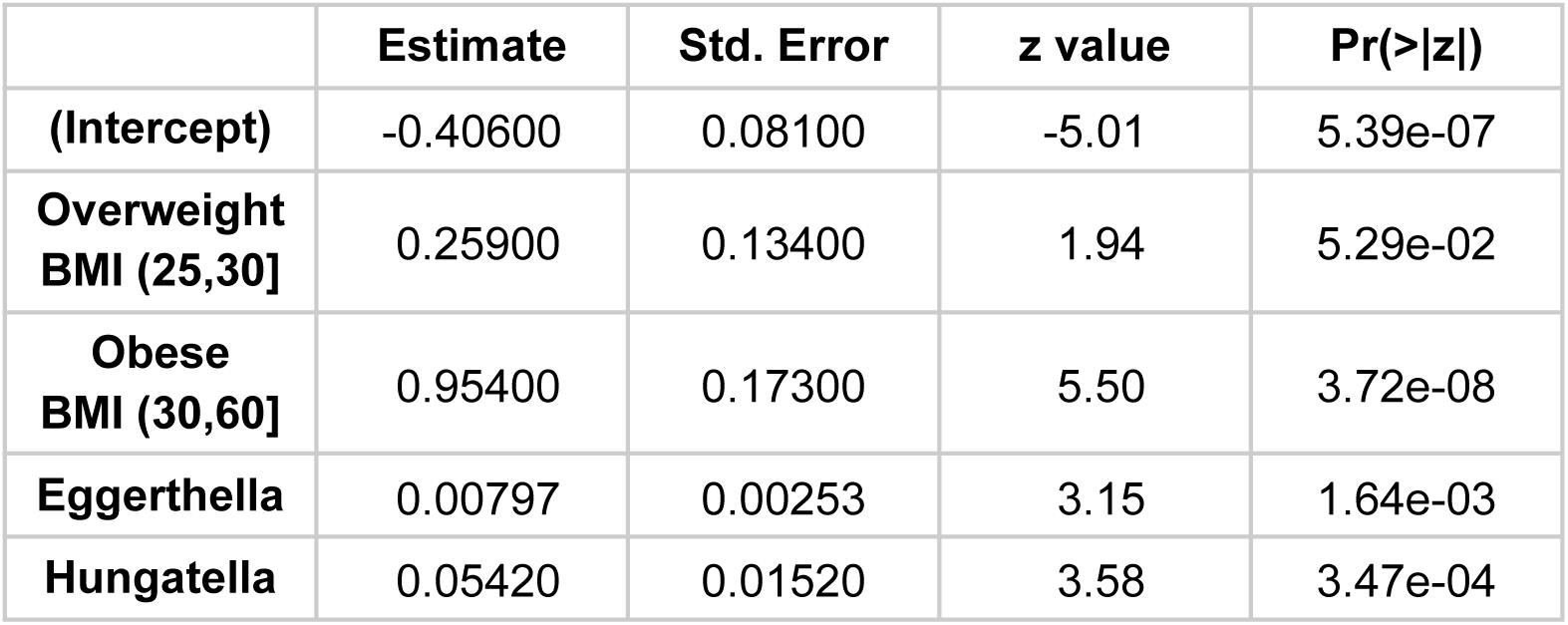
Coefficient estimates and their respective p-values of a logistic regression of MDD on *Eggerthella* and *Hungatella* abundances, and categorical BMI.

**sTable 8:** Causal effect of *Eggerthella* on MDD, expressed as the probability of experiencing MDD given a specific abundance level "e" of *Eggerthella* set by intervention (denoted as P(MDD=1|do(*Eggerthella*=e))), along with the corresponding 95% confidence interval.

| **Abundance of**<br>***Eggerthella*** | **Mean** | **95% Confidence Interval** |  |
| --- | --- | --- | --- |
| 0 | 0.472 | 0.426 | 0.520 |
| 24 | 0.520 | 0.471 | 0.568 |
| 46 | 0.562 | 0.498 | 0.625 |
| 70 | 0.608 | 0.522 | 0.688 |
| 92 | 0.649 | 0.541 | 0.743 |
| 116 | 0.690 | 0.560 | 0.796 |
| 138 | 0.726 | 0.577 | 0.837 |
| 162 | 0.762 | 0.595 | 0.874 |
| 184 | 0.792 | 0.611 | 0.902 |
| 208 | 0.821 | 0.628 | 0.926 |

**sTable 9:** Causal effect of Eggerthella on MDD, conditional on each category of BMI, expressed as the probability of experiencing MDD given a specific abundance level "e" of *Eggerthella* set by intervention and a specific BMI category "c" (denoted as P(MDD=1|do(*Eggerthella*=e), catBMI=c), along with the corresponding 95% confidence interval (C.I.).

| **Abundance of**<br>***Eggerthella*** | **Non- Obese** |  |  | **Overweight** |  |  | **Obese** |  |  |
| --- | --- | --- | --- | --- | --- | --- | --- | --- | --- |
|  | **Mean** | **95% C. I** |  | **Mean** | **95% C. I** |  | **Mean** | **95% C. I** |  |
| 0 | 0.421 | 0.378 | 0.465 | 0.482 | 0.421 | 0.544 | 0.652 | 0.574 | 0.723 |
| 24 | 0.468 | 0.422 | 0.515 | 0.530 | 0.468 | 0.591 | 0.694 | 0.620 | 0.760 |
| 46 | 0.512 | 0.449 | 0.575 | 0.574 | 0.500 | 0.644 | 0.730 | 0.652 | 0.796 |
| 70 | 0.559 | 0.472 | 0.643 | 0.620 | 0.527 | 0.704 | 0.766 | 0.679 | 0.835 |
| 92 | 0.602 | 0.492 | 0.703 | 0.660 | 0.549 | 0.756 | 0.796 | 0.700 | 0.867 |
| 116 | 0.647 | 0.512 | 0.762 | 0.701 | 0.570 | 0.806 | 0.825 | 0.719 | 0.897 |
| 138 | 0.686 | 0.530 | 0.809 | 0.737 | 0.588 | 0.846 | 0.849 | 0.735 | 0.920 |
| 162 | 0.725 | 0.548 | 0.852 | 0.772 | 0.607 | 0.881 | 0.872 | 0.751 | 0.939 |
| 184 | 0.759 | 0.566 | 0.884 | 0.802 | 0.624 | 0.908 | 0.890 | 0.765 | 0.953 |
| 208 | 0.792 | 0.584 | 0.912 | 0.830 | 0.642 | 0.930 | 0.908 | 0.779 | 0.965 |

**sTable 10:** Causal effect of *Hungatella* on MDD, expressed as the probability of experiencing MDD given a specific abundance level "e" of *Hungatella* set by intervention (denoted as P(MDD=1|do(*Hungatella*=e))), along with the corresponding 95% confidence interval.

| Abundance of *Hungatella* | Mean | 95% Confidence Interval |  |
| --- | --- | --- | --- |
| 0 | 0.474 | 0.428 | 0.521 |
| 6 | 0.557 | 0.499 | 0.614 |
| 11 | 0.624 | 0.539 | 0.702 |
| 17 | 0.698 | 0.581 | 0.794 |
| 22 | 0.753 | 0.614 | 0.854 |
| 28 | 0.809 | 0.651 | 0.906 |
| 33 | 0.848 | 0.68 | 0.936 |
| 39 | 0.886 | 0.713 | 0.961 |
| 44 | 0.911 | 0.739 | 0.974 |
| 50 | 0.935 | 0.767 | 0.984 |

**sTable 11:**
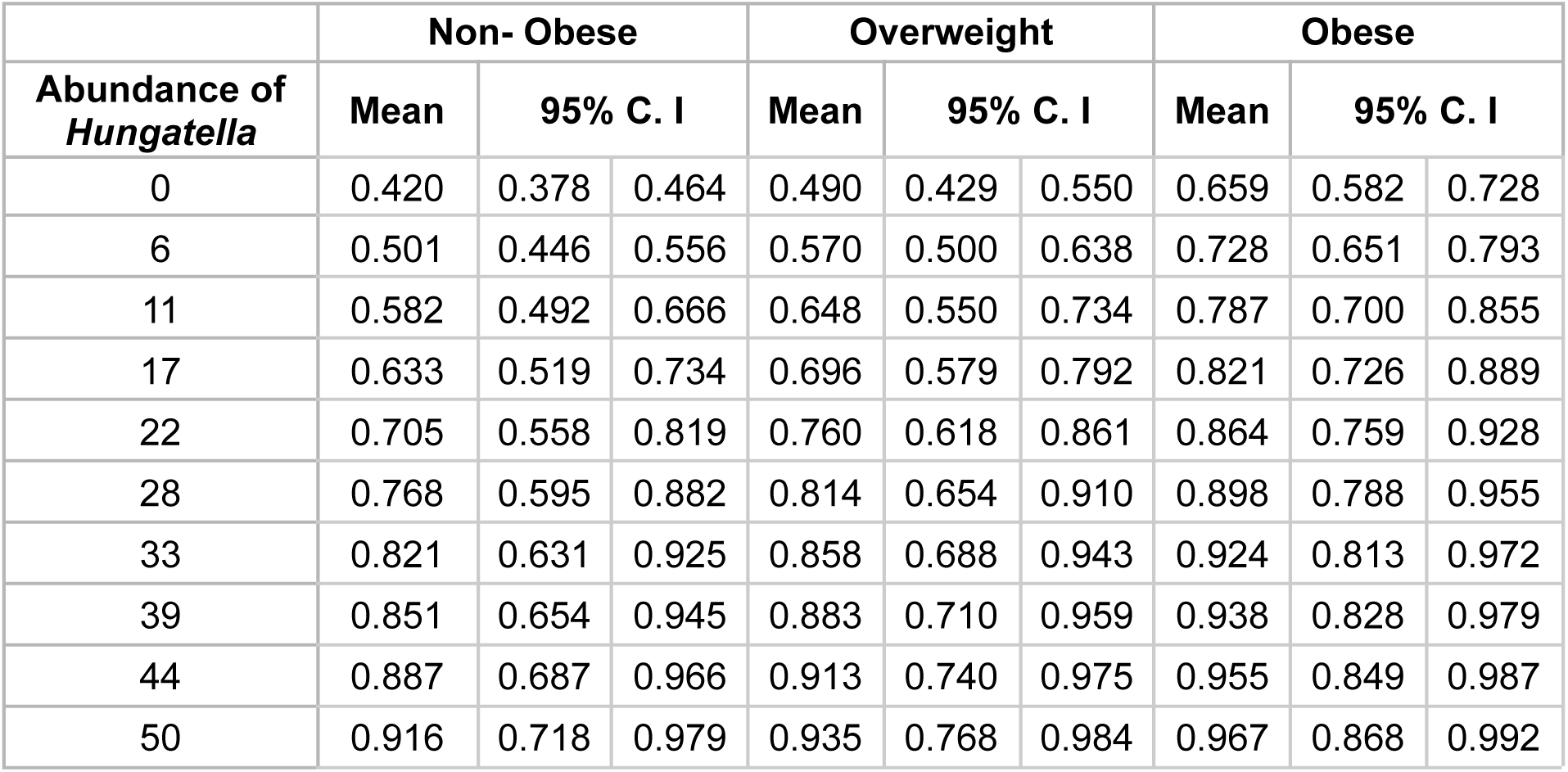
Causal effect of *Hungatella* on MDD, conditional on each category of BMI, expressed as the probability of experiencing MDD given a specific abundance level h" of *Hungatella* set by intervention and a specific BMI category "c" (denoted as P(MDD=1|do(*Hungatella*=h), catBMI=c), along with the corresponding 95% confidence interval (C.I.).

| Abundance of *Hungatella* | Non- Obese |  |  | Overweight |  |  | Obese |  |  |
| --- | --- | --- | --- | --- | --- | --- | --- | --- | --- |
|  | Mean | 95% C. I |  | Mean | 95% C. I |  | Mean | 95% C. I |  |
| 0 | 0.420 | 0.378 | 0.464 | 0.490 | 0.429 | 0.550 | 0.659 | 0.582 | 0.728 |
| 6 | 0.501 | 0.446 | 0.556 | 0.570 | 0.500 | 0.638 | 0.728 | 0.651 | 0.793 |
| 11 | 0.582 | 0.492 | 0.666 | 0.648 | 0.550 | 0.734 | 0.787 | 0.700 | 0.855 |
| 17 | 0.633 | 0.519 | 0.734 | 0.696 | 0.579 | 0.792 | 0.821 | 0.726 | 0.889 |
| 22 | 0.705 | 0.558 | 0.819 | 0.760 | 0.618 | 0.861 | 0.864 | 0.759 | 0.928 |
| 28 | 0.768 | 0.595 | 0.882 | 0.814 | 0.654 | 0.910 | 0.898 | 0.788 | 0.955 |
| 33 | 0.821 | 0.631 | 0.925 | 0.858 | 0.688 | 0.943 | 0.924 | 0.813 | 0.972 |
| 39 | 0.851 | 0.654 | 0.945 | 0.883 | 0.710 | 0.959 | 0.938 | 0.828 | 0.979 |
| 44 | 0.887 | 0.687 | 0.966 | 0.913 | 0.740 | 0.975 | 0.955 | 0.849 | 0.987 |
| 50 | 0.916 | 0.718 | 0.979 | 0.935 | 0.768 | 0.984 | 0.967 | 0.868 | 0.992 |

#### Effects of Discrete Abundances of Eggerthella and Hungatella on MDD

To enhance interpretability, we categorized Eggerthella abundance into two groups: ‘low’ (up to 30) and ‘high’ (exceeding 30). These groups demonstrate statistical significance at the 5% level, supported by a Pearson’s Chi-squared test with Yates’ continuity correction (X-squared = 19.458, df = 1, p-value = 1.028e-05). Similarly, *Hungatella* abundance was categorized into ‘low’ (up to 10) and ‘high’ (exceeding 10), showing statistical significance at the 5% level (X-squared = 11.09, df = 1, p-value = 0.0008678). These thresholds were chosen following an initial inspection of Figures 3 and 4, respectively. Further investigation is required to determine the optimal categorizations for both Eggerthella and Hungatella abundances.

#### Effects of Discrete Abundances of *Eggerthella* on MDD

Based on the discrete variables for Eggerthella abundance and BMI category, we can easily compute the post-interventional distributions of MDD for both levels of *Eggerthella*, and respective 95% confidence intervals. Specifically, P(MDD=1|do(*Eggerthella*=Low) = 0.4696, with 95% CI [0.4399, 0.4994]. In addition, P(MDD=1|do(*Eggerthella*=High) = 0.6703, with 95% CI [0.5911, 0.7408]. The Average Treatment Effect, expressed as ATE = E[MDD=1|do(*Eggerthella*=High)] - E[MDD=1|do(*Eggerthella*=Low)] = 0.168 (std. error 0.042) and a 95% Confidence Interval of (0.086, 0.250). Importantly, this difference is significantly non-zero.

We can also compute the post-interventional distributions of MDD for both low and high levels of *Eggerthella*, conditional on a specific value "c" of BMI category. *Supplementary Table 12* shows the estimates of P(MDD=1|do(*Eggerthella*=e), catBMI=c), for e = {Low, High}, while *Supplementary Figure 4* provides a visual representation of these probabilities. The causal effect sizes of Eggerthella at each "c" category of BMI, represented by the difference E(MDD=1|do(*Eggerthella*=High), catBMI=c) - E(MDD=1|do(*Eggerthella*=Low), catBMI=c), along with their respective standard errors and 95% confidence intervals, are detailed in *Supplementary Table 13*. Importantly, all effects are significantly distinct from zero.

These probabilities and effect sizes were all computed based on the estimates of P(MDD=1|*Eggerthela*=e, *Hungatella*=h, catBMI=c), modeled through a logistic regression. The coefficient estimates and their respective p-values are detailed in *Supplementary Table 14*.

**sTable 12:** Estimates of P(MDD=1|do(*Eggerthella*=e), catBMI=c), for both levels of *Eggerthella e* (Low or High), across the three categories of BMI, along with their respective 95% confidence intervals.

| BMI Category | Eggerthella | Mean | 95% CI |  |
| --- | --- | --- | --- | --- |
| **Non-Obese BMI (14,25]** | Low [0,30] | 0.4215 | 0.3853 | 0.4586 |
|  | High (30,209] | 0.6122 | 0.5251 | 0.6926 |
| **Overweight BMI (25,30]** | Low [0,30] | 0.4784 | 0.4234 | 0.5339 |
|  | High (30,209] | 0.6717 | 0.5826 | 0.7500 |
| **Obese BMI (30,60]** | Low [0,30] | 0.6511 | 0.5781 | 0.7177 |
|  | High (30,209] | 0.8012 | 0.7211 | 0.8627 |

**sFigure 4:**
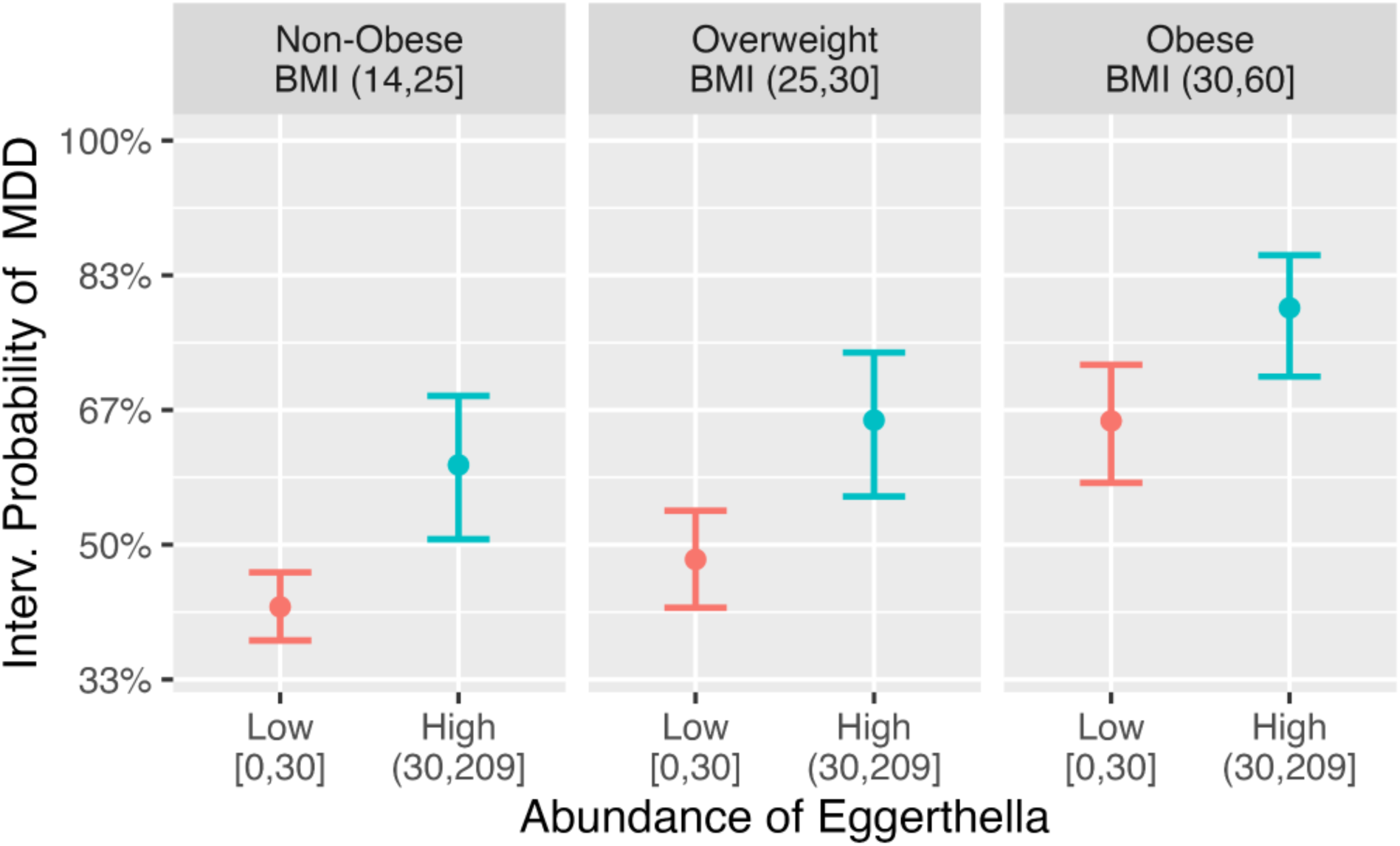
Estimated conditional post-interventional probabilities of MDD given *Eggerthella* levels. Estimated conditional post-interventional probabilities of MDD, given low and high levels of *Eggerthella* set by intervention, across the three categories of BMI, i.e., P(MDD=1|do(*Eggerthella*=Low), catBMI=c) and P(MDD=1|do(*Eggerthella*=High), catBMI=c), for each category of BMI "c". Error bars indicate the corresponding 95% confidence intervals.

**sTable 13:** Estimates of the causal effect sizes of *Eggerthella* at each category of BMI "c", represented by the difference E(MDD=1|do(*Eggerthella*=High), catBMI=c) - E(MDD=1|do(*Eggerthella*=Low), catBMI=c), along with their respective standard errors and 95% confidence intervals.

| BMI Category | Effect Size | Std. Error | 95% CI |  |
| --- | --- | --- | --- | --- |
| **No Obesity**<br>**BMI (14,25]** | 0.1742 | 0.0442 | 0.0875 | 0.2609 |
| **Overweight**<br>**BMI (25,30]** | 0.1712 | 0.0423 | 0.0883 | 0.2541 |
| **Obese**<br>**BMI (30,60]** | 0.1397 | 0.0330 | 0.0749 | 0.2045 |

**sTable 14:** Coefficient estimates and their respective p-values of a logistic regression of MDD on binary variables for the binary variables representing the abundances of *Eggerthella* and *Hungatella*, along with the variable indicating the category of BMI.

|  | Estimate | Std. Error | z value | Pr(>\|z\|) |
| --- | --- | --- | --- | --- |
| **(Intercept)** | -0.3537 | 0.0777 | -4.551 | 5.33e-06 |
| **Eggerthella -**<br>**High (30,191]** | 0.7099 | 0.1846 | 3.846 | 1.20e-04 |
| **Hungatella -**<br>**High (10,31]** | 0.8568 | 0.2860 | 2.996 | 2.74e-03 |
| **Overweight**<br>**BMI (25,30]** | 0.2372 | 0.1337 | 1.774 | 7.60e-02 |
| **Obese**<br>**BMI (30,60]** | 0.9458 | 0.1732 | 5.460 | 4.76e-08 |

#### Effects of Discrete Abundances of *Hungatella* on MDD

Based on the discrete variables for *Eggerthella* abundance and BMI category, we also computed the post-interventional distributions of MDD for both levels of *Hungatella*, and respective 95% confidence intervals. Specifically, P(MDD=1|do(*Hungatella*=Low) = 0.4824, with 95% CI [0.4537, 0.5112]. In addition, P(MDD=1|do(*Hungatella*=High) = 0.7130, with 95% CI [0.5897, 0.8111]. The ATE = E[MDD=1|do(Hungatella=High)] - E[MDD=1|do(Hungatella=Low)] = 0.1984 (std. error 0.0608) and a 95% confidence interval of (0.0792, 0.3176). Importantly, this difference is also significantly different from zero.

We can also compute the post-interventional distributions of MDD for both low and high levels of *Hungatella*, conditional on a specific category "c" of BMI. *Supplementary Table 15* shows the estimates of P(MDD=1|do(*Hungatella*=e), catBMI=c), for e = low, high, while *Supplementary Figure 5* provides a visual representation of these probabilities.

The causal effect sizes of *Hungatella* at each category "c" of BMI, represented by the difference E(MDD=1|do(*Hungatella*=High), catBMI=c) - E(MDD=1|do(*Hungatella*=Low), catBMI=c), along with their respective standard errors and 95% confidence intervals, are detailed in *Supplementary Table 16*. Importantly, all the differences are significantly distinct from zero.

Once again, all these estimates were computed based on the estimates of P(MDD=1|*Eggerthela*=e, *Hungatella*=h, catBMI=c), modeled through a logistic regression – see *Supplementary Table 14* for coefficient estimates and their respective p-values.

**sTable 15:** Estimates of P(MDD=1|do(*Hungatella*=h), catBMI=c), for both low and high levels of *Hungatella*, across the three categories of BMI, along with their respective 95% confidence intervals.

| BMI Category | *Hungatella* | $P(\text{MDD}=1\vert $<br>$\text{do}(\text{Hungatella}=h),$<br>$\text{catBMI}=c)$ | 95% CI | |
| --- | --- | --- | --- | --- |
| Non-Obese<br>BMI (14,25] | Low [0,10] | 0.4291 | 0.3932 | 0.4658 |
|  | High (10,50] | 0.6618 | 0.5297 | 0.7727 |
| Overweight<br>BMI (25,30] | Low [0,10] | 0.4944 | 0.4401 | 0.5488 |
|  | High (10,50] | 0.7374 | 0.6109 | 0.8340 |
| Obese<br>BMI (30,60] | Low [0,10] | 0.6661 | 0.5948 | 0.7306 |
|  | High (10,50] | 0.8436 | 0.7430 | 0.9097 |

**sFigure 5:**
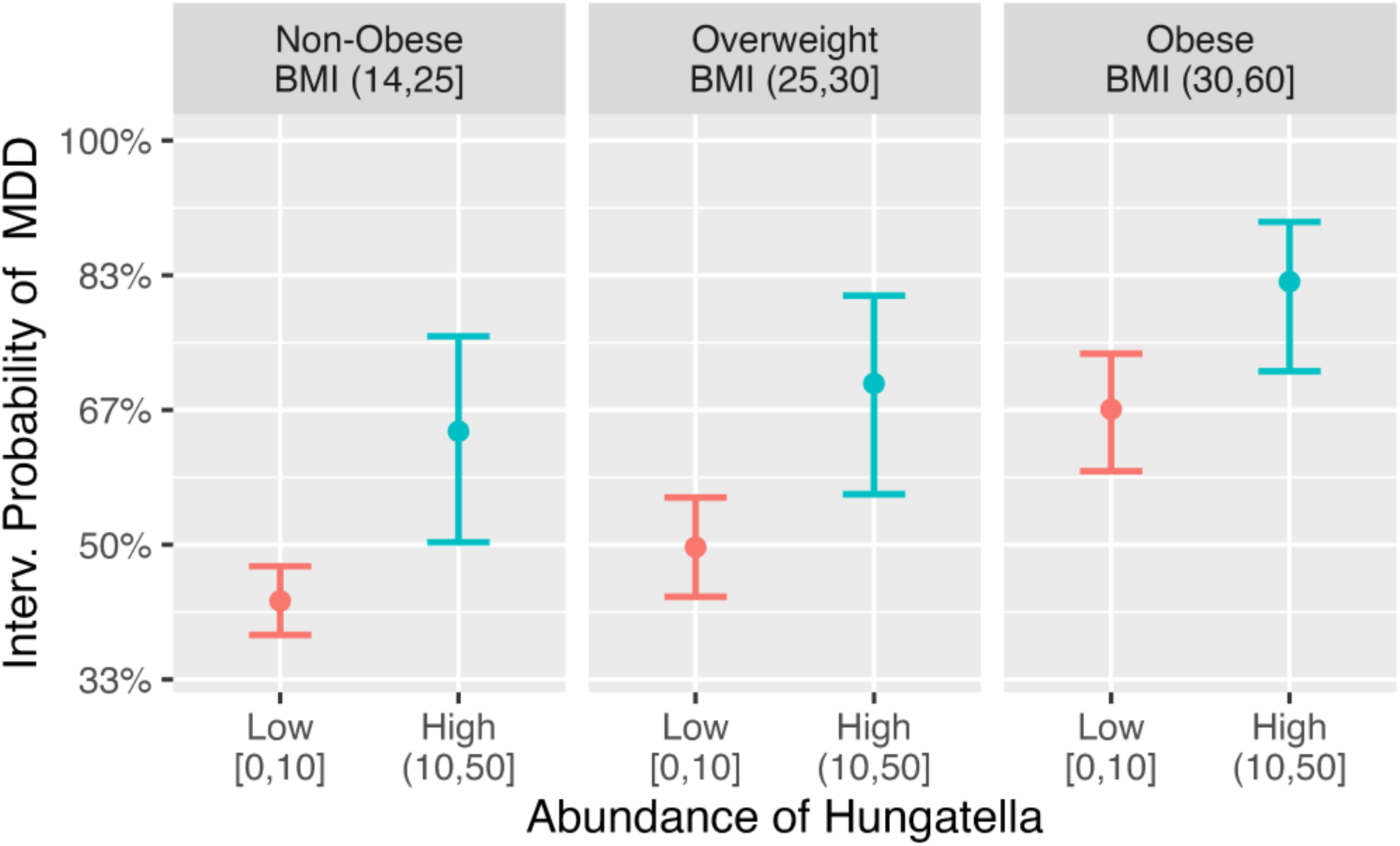
Estimated conditional post-interventional probabilities of MDD given *Hungatella* levels. Estimated conditional post-interventional probabilities of MDD, given low and high levels of *Hungatella* set by intervention, across the three categories of BMI, i.e., P(MDD=1|do(*Hungatella*=Low), catBMI=c) and P(MDD=1|do(*Hungatella*=High), catBMI=c), for each category of BMI "c". Error bars indicate the corresponding 95% confidence intervals.

**sTable 16:**
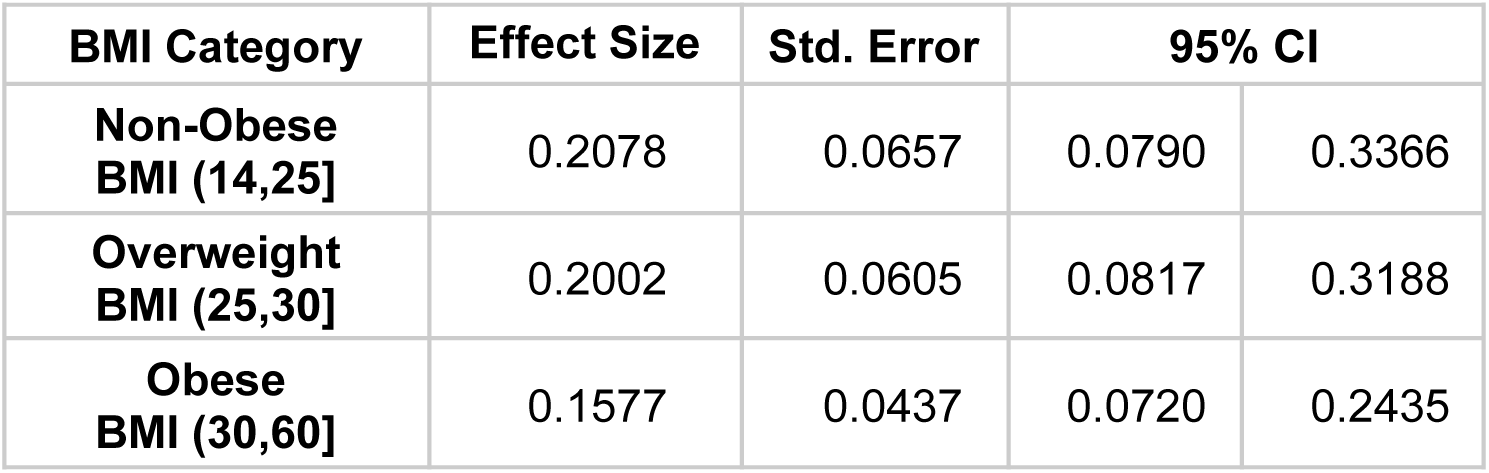
Estimates of the causal effect sizes of *Hungatella* at each category of BMI "c", represented by the difference E(MDD=1|do(*Hungatella*=High), catBMI=c) - E(MDD=1|do(*Hungatella*=Low), catBMI=c), along with their respective standard errors and 95% confidence intervals.

| BMI Category | Effect Size | Std. Error | 95% CI |  |
| --- | --- | --- | --- | --- |
| **Non-Obese BMI (14,25]** | 0.2078 | 0.0657 | 0.0790 | 0.3366 |
| **Overweight BMI (25,30]** | 0.2002 | 0.0605 | 0.0817 | 0.3188 |
| **Obese BMI (30,60]** | 0.1577 | 0.0437 | 0.0720 | 0.2435 |

#### Relative Risks of MDD Associated with High *Eggerthella* Levels

*Supplementary Table 17* presents the relative risks of MDD associated with high *Eggerthella* levels compared to low levels in each category of BMI. Additionally, it includes the respective standard errors and 95% confidence intervals. Notably, all relative risks are significantly greater than one. In particular, high levels of *Eggerthella* increase the risk of MDD by 42.6% (95% CI: 20.2 – 65.0%) among non-obese individuals, by 36.8% (95% CI: 18.0 – 55.5%) for individuals with overweight, and by 22.1% (95% CI: 11.7 – 32.5%) for those with Obesity.

**sTable 17:** Relative risks of MDD associated with high *Eggerthella* abundance levels compared to low abundance levels within each category of BMI.

| BMI Category | Relative Risk | Std. Error | 95% CI |  |
| --- | --- | --- | --- | --- |
| **Non-Obese**<br>**BMI (14,25]** | 1.426 | 0.11440 | 1.202 | 1.650 |
| **Overweight**<br>**BMI (25,30]** | 1.368 | 0.09569 | 1.180 | 1.555 |
| **Obese**<br>**BMI (30,60]** | 1.221 | 0.05289 | 1.117 | 1.325 |

#### Relative Risks of MDD Associated with High *Hungatella* Levels

*Supplementary Table 18* presents the relative risks of MDD associated with high *Hungatella* levels compared to low levels in each category of BMI. Additionally, it includes the respective standard errors and 95% confidence intervals. All relative risks are significantly greater than one. In particular, high levels of *Hungatella* increase the risk of MDD by 51.1% (95% CI: 18.1 – 84.1%) among non-obese individuals, by 43.8% (95% CI: 16.7 – 70.9%) for individuals with overweight, and by 25.8% (95% CI: 11.5 – 40.0%) for those with Obesity..

**sTable 18:** Relative risks of MDD associated with high *Hungatella* abundance levels compared to low abundance levels within each category of BMI.

| BMI Category | Relative Risk | Std. Error | 95% CI |  |
| --- | --- | --- | --- | --- |
| **Non-Obese**<br>**BMI (14,25]** | 1.511 | 0.16840 | 1.181 | 1.841 |
| **Overweight**<br>**BMI (25,30]** | 1.438 | 0.13820 | 1.167 | 1.709 |
| **Obese**<br>**BMI (30,60]** | 1.258 | 0.07267 | 1.115 | 1.400 |

## References

1. GBD 2019 Mental Disorders Collaborators. Global, regional, and national burden of 12 mental disorders in 204 countries and territories, 1990–2019: a systematic analysis for the Global Burden of Disease Study 2019. Lancet Psychiatry. 2022;9(2):137–150. doi:10.1016/s2215-0366(21)00395-3

2. Winter NR, Blanke J, Leenings R, et al. A Systematic Evaluation of Machine Learning–Based Biomarkers for Major Depressive Disorder. JAMA Psychiatry. 2024;81(4). doi:10.1001/jamapsychiatry.2023.5083

3. Winter NR, Leenings R, Ernsting J, et al. Quantifying Deviations of Brain Structure and Function in Major Depressive Disorder Across Neuroimaging Modalities. JAMA Psychiatry. 2022;79(9):879–888. doi:10.1001/jamapsychiatry.2022.1780

4. Flint J. The genetic basis of major depressive disorder. Mol Psychiatry. 2023;28(6):2254–2265. doi:10.1038/s41380-023-01957-9

5. Malhi GS, Mann JJ. Depression. The Lancet. 2018;392(10161):2299–2312. doi:10.1016/s0140-6736(18)31948-2

6. Kelly JR, Borre Y, O’ Brien C, et al. Transferring the blues: Depression-associated gut microbiota induces neurobehavioural changes in the rat. J Psychiatr Res. 2016;82:109–118. doi:10.1016/j.jpsychires.2016.07.019

7. Cryan JF, O’Riordan KJ, Cowan CSM, et al. The Microbiota-Gut-Brain Axis. Physiol Rev. 2019;99(4):1877–2013. doi:10.1152/physrev.00018.2018

8. Gao M, Wang J, Liu P, et al. Gut microbiota composition in depressive disorder: a systematic review, meta-analysis, and meta-regression. Transl Psychiatry. 2023;13(1):379. doi:10.1038/s41398-023-02670-5

9. Nikolova VL, Smith MRB, Hall LJ, Cleare AJ, Stone JM, Young AH. Perturbations in Gut Microbiota Composition in Psychiatric Disorders: A Review and Meta-analysis. JAMA Psychiatry. 2021;78(12):1343. doi:10.1001/jamapsychiatry.2021.2573

10. Łoniewski I, Misera A, Skonieczna-Żydecka K, et al. Major Depressive Disorder and gut microbiota – Association not causation. A scoping review. Prog Neuropsychopharmacol Biol Psychiatry. 2021;106:110111. doi:10.1016/j.pnpbp.2020.110111

11. Bruce-Keller AJ, Salbaum JM, Luo M, et al. Obese-type Gut Microbiota Induce Neurobehavioral Changes in the Absence of Obesity. Biol Psychiatry. 2015;77(7):607–615. doi:10.1016/j.biopsych.2014.07.012

12. Radjabzadeh D, Bosch JA, Uitterlinden AG, et al. Gut microbiome-wide association study of depressive symptoms. Nat Commun. 2022;13(1):7128. doi:10.1038/s41467-022-34502-3

13. Davey Smith G, Hemani G. Mendelian randomization: genetic anchors for causal inference in epidemiological studies. Hum Mol Genet. 2014;23(R1):R89–R98. doi:10.1093/hmg/ddu328

14. Ribeiro AH, Soler JMP, Neto EC, Fujita A. Causal Inference and Structure Learning of Genotype–Phenotype Networks Using Genetic Variation. In: Wong KC, ed. Big Data Analytics in Genomics. Springer International Publishing; 2016:89–143. doi:10.1007/978-3-319-41279-5_3

15. Feuerriegel S, Frauen D, Melnychuk V, et al. Causal machine learning for predicting treatment outcomes. Nat Med. 2024;30(4):958–968. doi:10.1038/s41591-024-02902-1

16. de Leeuw C, Savage J, Bucur IG, Heskes T, Posthuma D. Understanding the assumptions underlying Mendelian randomization. Eur J Hum Genet. 2022;30(6):653–660. doi:10.1038/s41431-022-01038-5

17. Sekula P, Del Greco M F, Pattaro C, Köttgen A. Mendelian Randomization as an Approach to Assess Causality Using Observational Data. J Am Soc Nephrol JASN. 2016;27(11):3253–3265. doi:10.1681/ASN.2016010098

18. Vogelbacher C, Möbius TWD, Sommer J, et al. The Marburg-Münster Affective Disorders Cohort Study (MACS): A quality assurance protocol for MR neuroimaging data. NeuroImage. 2018;172:450–460. doi:10.1016/j.neuroimage.2018.01.079

19. Pearl J. Causality: Models, Reasoning and Inference by Judea Pearl(2009-09-14). Cambridge University Press; 2009.

20. Zhang J. On the completeness of orientation rules for causal discovery in the presence of latent confounders and selection bias. Artif Intell. 2008;172(16):1873–1896. doi:10.1016/j.artint.2008.08.001

21. Perković E, Textor J, Kalisch M, Maathuis MH. Complete Graphical Characterization and Construction of Adjustment Sets in Markov Equivalence Classes of Ancestral Graphs. J Mach Learn Res. Published online May 2018:62 p. doi:10.3929/ETHZ-B-000278021

22. Jaber A, Ribeiro A, Zhang J, Bareinboim E. Causal Identification under Markov equivalence: Calculus, Algorithm, and Completeness. Adv Neural Inf Process Syst. 2022;35:3679–3690.

23. Pearl J. Myth, Confusion, and Science in Causal Analysis. Technichal Report. University of California, Los Angeles; 2009. https://ucla.in/2EihVyD

24. Pearl J. Invited Commentary: Understanding Bias Amplification. Am J Epidemiol. 2011;174(11):1223–1227. doi:10.1093/aje/kwr352

25. Wittchen HU, Wunderlich U, Gruschwitz S, Zaudig M. SKID I. Strukturiertes Klinisches Interview für DSM-IV. Achse I: Psychische Störungen. Interviewheft und Beurteilungsheft. Eine deutschsprachige, erweiterte Bearb. d. amerikanischen Originalversion des SKID I. Published online 1997. Accessed November 26, 2024. https://hdl.handle.net/11858/00-001M-0000-000E-AAB1-4

26. Troci A, Zimmermann O, Esser D, et al. B-cell-depletion reverses dysbiosis of the microbiome in multiple sclerosis patients. Sci Rep. 2022;12(1):3728. doi:10.1038/s41598-022-07336-8

27. Trautmann T, Bang C, Franke A, Vincent D, Reinshagen K, Boettcher M. The Impact of Oral Sodium Chloride Supplementation on Thrive and the Intestinal Microbiome in Neonates With Small Bowel Ostomies: A Prospective Cohort Study. Front Immunol. 2020;11:1421. doi:10.3389/fimmu.2020.01421

28. Caporaso JG, Lauber CL, Walters WA, et al. Ultra-high-throughput microbial community analysis on the Illumina HiSeq and MiSeq platforms. ISME J. 2012;6(8):1621–1624. doi:10.1038/ismej.2012.8

29. Welzel M, Lange A, Heider D, et al. Natrix: a Snakemake-based workflow for processing, clustering, and taxonomically assigning amplicon sequencing reads. BMC Bioinformatics. 2020;21(1):526. doi:10.1186/s12859-020-03852-4

30. Köster J, Rahmann S. Snakemake—a scalable bioinformatics workflow engine. Bioinformatics. 2012;28(19):2520–2522. doi:10.1093/bioinformatics/bts480

31. Schmieder R, Edwards R. Quality control and preprocessing of metagenomic datasets. Bioinformatics. 2011;27(6):863–864. doi:10.1093/bioinformatics/btr026

32. Masella AP, Bartram AK, Truszkowski JM, Brown DG, Neufeld JD. PANDAseq: paired-end assembler for illumina sequences. BMC Bioinformatics. 2012;13(1):31. doi:10.1186/1471-2105-13-31

33. Fu L, Niu B, Zhu Z, Wu S, Li W. CD-HIT: accelerated for clustering the next-generation sequencing data. Bioinformatics. 2012;28(23):3150–3152. doi:10.1093/bioinformatics/bts565

34. Rognes T, Flouri T, Nichols B, Quince C, Mahé F. VSEARCH: a versatile open source tool for metagenomics. PeerJ. 2016;4:e2584. doi:10.7717/peerj.2584

35. Mahé F, Rognes T, Quince C, Vargas C de, Dunthorn M. Swarm: robust and fast clustering method for amplicon-based studies. PeerJ. 2014;2:e593. doi:10.7717/peerj.593

36. Camacho C, Coulouris G, Avagyan V, et al. BLAST+: architecture and applications. BMC Bioinformatics. 2009;10(1):421. doi:10.1186/1471-2105-10-421

37. Quast C, Pruesse E, Yilmaz P, et al. The SILVA ribosomal RNA gene database project: improved data processing and web-based tools. Nucleic Acids Res. 2013;41(D1):D590–D596. doi:10.1093/nar/gks1219

38. Johnson JS, Spakowicz DJ, Hong BY, et al. Evaluation of 16S rRNA gene sequencing for species and strain-level microbiome analysis. Nat Commun. 2019;10(1):5029. doi:10.1038/s41467-019-13036-1

39. Yang L, Chen J. A comprehensive evaluation of microbial differential abundance analysis methods: current status and potential solutions. Microbiome. 2022;10(1):130. doi:10.1186/s40168-022-01320-0

40. Zhou H, He K, Chen J, Zhang X. LinDA: linear models for differential abundance analysis of microbiome compositional data. Genome Biol. 2022;23(1):95. doi:10.1186/s13059-022-02655-5

41. Spirtes P, Glymour C, Scheines R. Causation, Prediction, and Search. The MIT Press; 2001. doi:10.7551/mitpress/1754.001.0001

42. Mooij JM, Claassen T. Constraint-Based Causal Discovery using Partial Ancestral Graphs in the presence of Cycles. In: Proceedings of the 36th Conference on Uncertainty in Artificial Intelligence (UAI). PMLR; 2020:1159–1168. Accessed October 17, 2024. https://proceedings.mlr.press/v124/m-mooij20a.html

43. Kalisch M, Mächler M, Colombo D, Maathuis MH, Bühlmann P. Causal Inference Using Graphical Models with the R Package pcalg. J Stat Softw. 2012;47:1–26. doi:10.18637/jss.v047.i11

44. Tsagris M, Borboudakis G, Lagani V, Tsamardinos I. Constraint-based causal discovery with mixed data. Int J Data Sci Anal. 2018;6(1):19–30. doi:10.1007/s41060-018-0097-y

45. Lagani V, Athineou G, Farcomeni A, Tsagris M, Tsamardinos I. Feature Selection with the R Package MXM: Discovering Statistically Equivalent Feature Subsets. J Stat Softw. 2017;80:1–25. doi:10.18637/jss.v080.i07

46. Roumpelaki A, Borboudakis G, Triantafillou S, Tsamardinos I. Marginal causal consistency in constraint-based causal learning. In: In Causation: Foundation to Application Workshop, UAI.; 2016.

47. Colombo D, Maathuis MH, Kalisch M, Richardson TS. Learning high-dimensional directed acyclic graphs with latent and selection variables. Ann Stat. 2012;40(1):294–321. doi:10.1214/11-AOS940

48. Maathuis MH, Colombo D. A generalized back-door criterion. Ann Stat. 2015;43(3):1060–1088. doi:10.1214/14-AOS1295

49. Knudsen JK, Michaelsen TY, Bundgaard-Nielsen C, et al. Faecal microbiota transplantation from patients with depression or healthy individuals into rats modulates mood-related behaviour. Sci Rep. 2021;11(1):21869. doi:10.1038/s41598-021-01248-9

50. Cekanaviciute E, Yoo BB, Runia TF, et al. Gut bacteria from multiple sclerosis patients modulate human T cells and exacerbate symptoms in mouse models. Proc Natl Acad Sci. 2017;114(40):10713–10718. doi:10.1073/pnas.1711235114

51. Wang Q, Li F, Liang B, et al. A metagenome-wide association study of gut microbiota in asthma in UK adults. BMC Microbiol. 2018;18(1):114. doi:10.1186/s12866-018-1257-x

52. Balakrishnan B, Luckey D, Wright K, Davis JM, Chen J, Taneja V. Eggerthella lenta augments preclinical autoantibody production and metabolic shift mimicking senescence in arthritis. Sci Adv. 2023;9(35):eadg1129. doi:10.1126/sciadv.adg1129

53. Müller B, Rasmusson AJ, Just D, et al. Fecal Short-Chain Fatty Acid Ratios as Related to Gastrointestinal and Depressive Symptoms in Young Adults. Psychosom Med. 2021;83(7):693–699. doi:10.1097/PSY.0000000000000965

54. Ma S, Xie X, Deng Z, et al. A Machine Learning Analysis of Big Metabolomics Data for Classifying Depression: Model Development and Validation. Biol Psychiatry. 2024;96(1):44–56. doi:10.1016/j.biopsych.2023.12.015

55. Milaneschi Y, Lamers F, Berk M, Penninx BWJH. Depression Heterogeneity and Its Biological Underpinnings: Toward Immunometabolic Depression. Biol Psychiatry. 2020;88(5):369–380. doi:10.1016/j.biopsych.2020.01.014

56. Kaur S, Yawar M, Kumar PA, Suresh K. Hungatella effluvii gen. nov., sp. nov., an obligately anaerobic bacterium isolated from an effluent treatment plant, and reclassification of Clostridium hathewayi as Hungatella hathewayi gen. nov., comb. nov. Int J Syst Evol Microbiol. 2014;64(Pt_3):710–718. doi:10.1099/ijs.0.056986-0

57. Chan CWH, Leung TF, Choi KC, et al. Association of early-life gut microbiome and lifestyle factors in the development of eczema in Hong Kong infants. Exp Dermatol. 2021;30(6):859–864. doi:10.1111/exd.14280

58. Li N, Bai C, Zhao L, Sun Z, Ge Y, Li X. The Relationship Between Gut Microbiome Features and Chemotherapy Response in Gastrointestinal Cancer. Front Oncol. 2021;11:781697. doi:10.3389/fonc.2021.781697

59. Misiak B, Pawlak E, Rembacz K, et al. Associations of gut microbiota alterations with clinical, metabolic, and immune-inflammatory characteristics of chronic schizophrenia. J Psychiatr Res. 2024;171:152–160. doi:10.1016/j.jpsychires.2024.01.036

60. Jantaratnotai N, Mosikanon K, Lee Y, McIntyre RS. The interface of depression and obesity. Obes Res Clin Pract. 2017;11(1):1–10. doi:10.1016/j.orcp.2016.07.003

61. Castaner O, Goday A, Park YM, et al. The Gut Microbiome Profile in Obesity: A Systematic Review. Int J Endocrinol. 2018;2018:1–9. doi:10.1155/2018/4095789

62. Ait Chait Y, Mottawea W, Tompkins TA, Hammami R. Unravelling the antimicrobial action of antidepressants on gut commensal microbes. Sci Rep. 2020;10(1):17878. doi:10.1038/s41598-020-74934-9

63. Macedo D, Filho AJMC, Soares de Sousa CN, et al. Antidepressants, antimicrobials or both? Gut microbiota dysbiosis in depression and possible implications of the antimicrobial effects of antidepressant drugs for antidepressant effectiveness. J Affect Disord. 2017;208:22–32. doi:10.1016/j.jad.2016.09.012

64. Colombo D, Maathuis MH. Order-independent constraint-based causal structure learning. J Mach Learn Res. 2014;15(1):3741–3782.

65. Zhang J, Spirtes P. Detection of Unfaithfulness and Robust Causal Inference. Minds Mach. 2008;18(2):239–271. doi:10.1007/s11023-008-9096-4

66. Zhalama, Zhang J, Mayer W. Weakening faithfulness: some heuristic causal discovery algorithms. Int J Data Sci Anal. 2017;3(2):93–104. doi:10.1007/s41060-016-0033-y

